# Lung proteomic biomarkers associated with chronic obstructive pulmonary disease

**DOI:** 10.1101/2021.04.07.21255030

**Authors:** Yu-Hang Zhang, Michael R. Hoopmann, Peter J. Castaldi, Kirsten A. Simonsen, Mukul K. Midha, Michael H. Cho, Gerard J. Criner, Raphael Bueno, Jiangyuan Liu, Robert L. Moritz, Edwin K. Silverman

## Abstract

**Background:** Identifying protein biomarkers for chronic obstructive pulmonary disease (COPD) has been challenging. Most previous studies have utilized individual proteins or pre-selected protein panels measured in blood samples. Mass spectrometry proteomic studies of lung tissue have been based on small sample sizes.

**Methods:** We utilized mass spectrometry proteomic approaches to discover protein biomarkers from 150 lung tissue samples representing COPD cases and controls. Top COPD-associated proteins were identified based on multiple linear regression analysis with false discovery rate (FDR) < 0.05. Correlations between pairs of COPD-associated proteins were examined. Machine learning models were also evaluated to identify potential combinations of protein biomarkers related to COPD.

**Results:** We identified 4407 proteins passing quality controls. Twenty-five proteins were significantly associated with COPD at FDR < 0.05, including Interleukin 33, Ferritin (light chain and heavy chain), and two proteins related to caveolae (CAV1 and CAVIN1). Multiple previously reported plasma protein biomarkers for COPD were not significantly associated with proteomic analysis of COPD in lung tissue, although RAGE was borderline significant. Eleven pairs of top significant proteins were highly correlated (r > 0.8), including several strongly correlated with RAGE (EHD2 and CAVIN1). Machine learning models using Random Forests with the top 5% of protein biomarkers demonstrated reasonable accuracy (0.766) and AUC (0.702) for COPD prediction.

**Conclusions:** Mass spectrometry proteomic analysis of lung tissue is a promising approach for the identification of biomarkers for COPD.

## Introduction

Chronic obstructive pulmonary disease (COPD), a major public health problem diagnosed by persistent airflow obstruction, is associated with chronic lung inflammation that can persist decades after smoking cessation (McDonough et al., 2011). In the United States, COPD is the fourth leading cause of death (Kochanek, Murphy, Xu, & Arias, 2017) and affects more than 16 million adults (Sullivan et al., 2018). COPD is typically diagnosed using spirometry; however, chronic airflow obstruction results from a heterogeneous combination of emphysema, small airway disease, and large airway disease (Castaldi et al., 2020). Molecular biomarkers could potentially assist in COPD diagnosis. COPD also has variable rates of onset and progression, and molecular biomarkers could identify individuals at high risk for disease development or progression, who may be candidates for more aggressive therapeutic interventions (Andreeva et al., 2017; Foong & Hall, 2016; Paone et al., 2016).

To identify molecular biomarkers for COPD pathogenesis and/or progression, various Omics data types have been used, including transcriptomics, epigenetics, metabolomics, and proteomics (Fortis et al., 2019). As key molecular agents, proteins are of particular interest as potential disease biomarkers. Previous studies have been performed using both single and multiple protein biomarkers. Single protein biomarkers associated with COPD have included SFTPD (Surfactant Protein D, encoded by *SFTPD*) (D. A. Lomas et al., 2009), Fibrinogen (encoded by *FGA, FGB* and *FGG*) (Danesh et al., 2005; Groenewegen et al., 2008), CC-16 (Club cell secretory protein-16, encoded by *SCGB1A1*) (David A Lomas et al., 2008), sRAGE (Advanced Glycosylation End-Product Specific Receptor, encoded by *AGER*) (Smith et al., 2011) and IL-6 (Interleukin 6, encoded by *IL6*) (Hurst et al., 2006).

Protein biomarker panels measured in blood samples have also been investigated in COPD. In 2007, a panel of 24 serum protein biomarkers was shown to be correlated with important clinical outcomes of COPD including forced expiratory volume in 1 s (FEV_1_), carbon monoxide transfer factor, 6-minute walk distance, and exacerbation frequency (Pinto-Plata et al., 2007). In 2011, four plasma biomarkers (α2-macroglobulin, haptoglobin, ceruloplasmin, and hemopexin) were able to distinguish patients with asthma, COPD and normal controls (Verrills et al., 2011). More recently, larger panels of protein biomarkers have been associated with COPD severity (Zemans et al., 2017) and exacerbations (Keene et al., 2017). However, most previous protein biomarker studies in COPD have utilized pre-selected protein biomarkers rather than unbiased assessments of all available proteins, and they have focused on blood samples rather than lung samples. A smaller number of studies have used mass spectrometry approaches for large-scale proteomic assessments in lung tissue samples, but their sample sizes have been limited (Brandsma et al., 2020; Cornwell et al., 2019; E. J. Lee et al., 2009).

We hypothesized that mass spectrometry analysis of lung tissue samples in COPD cases and smokers with normal spirometry would identify novel protein biomarkers for COPD. We assessed association of individual proteins with COPD case-control status, and we utilized machine learning methods to identify combinations of proteins associated with COPD. We also assessed the correlations between top protein biomarkers to understand potential biological relationships, and we compared our lung tissue protein biomarkers to previously reported COPD protein biomarkers. The integrated analyses not only identified new candidate COPD proteomic biomarkers, but also provided an initial step towards building predictive models for COPD.

## Methods

Detailed materials and methods are available in the Supplemental Materials, including the overall workflow (**Supplemental Figure S1**).

### Study Population

We analyzed lung tissue samples from ex-smokers obtained during clinical thoracic surgery procedures. Most samples were obtained from the NHLBI Lung Tissue Research Consortium (n=109, with 83 COPD cases and 26 control smokers). In addition, 43 subjects from a previously reported lung tissue population were also included (17 COPD cases and 26 control smokers) (Morrow et al., 2017).

### Sample Preparation and Mass Spectrometry Analysis

Proteins were extracted from lung tissue through mechanical shearing followed by cryolysis. Each sample was analyzed in triplicate by high resolution nano liquid chromatography with tandem mass spectrometry (LC-MS/MS). The raw mass spectra output files were analyzed using the Trans-Proteomic Pipeline (Deutsch et al., 2015). The final protein quantity was estimated as the sum of up to the top three most abundant peptides among the proteotypic peptides (i.e., peptides that match to only a single protein), using the same peptides for each protein in each sample.

### Biomarker Identification

Proteins with > 50% missing values and two outlier samples based on missingness were removed (**Supplemental Figure S2**). Principal component analyses were used to assess the data distribution before and after data preprocessing steps. After normalization, imputation, and batch effect correction, linear regression analyses were applied to identify COPD-associated proteomic biomarkers:

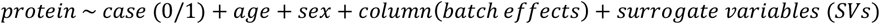

The False Discovery Rate (FDR) was controlled at 5%. Additional linear regression models with covariates including body mass index (BMI), pack-years of smoking, and lung cancer status were also assessed (see **Supplemental Materials Tables S1-S8)**. Pearson correlation analysis was performed to assess the correlations between the top 5% of proteins associated with COPD.

### Machine Learning Analysis

Using the top 5% of proteins associated with COPD, we performed machine learning analysis using Random Forests, Naïve Bayes, and Elastic Net with five-fold cross-validation. For each machine learning model, the averaged accuracy, area under the curve in receiver-operator curve analysis (AUC), and feature importance were compared.

### Functional Enrichment Analysis

Gene ontology (GO) enrichment analysis using the *topGO* algorithm was applied to the top 5% of proteins generated from the linear regression analyses to investigate potential related biological functions. Additional GO enrichment analyses with the same parameters were also applied to the top 5% of proteins positively and negatively associated with COPD.

## Results

### Clinical characteristics

We included 152 lung tissue samples in our proteomics analysis. The clinical characteristics of these subjects are shown in **Table 1**.

**Table 1:**
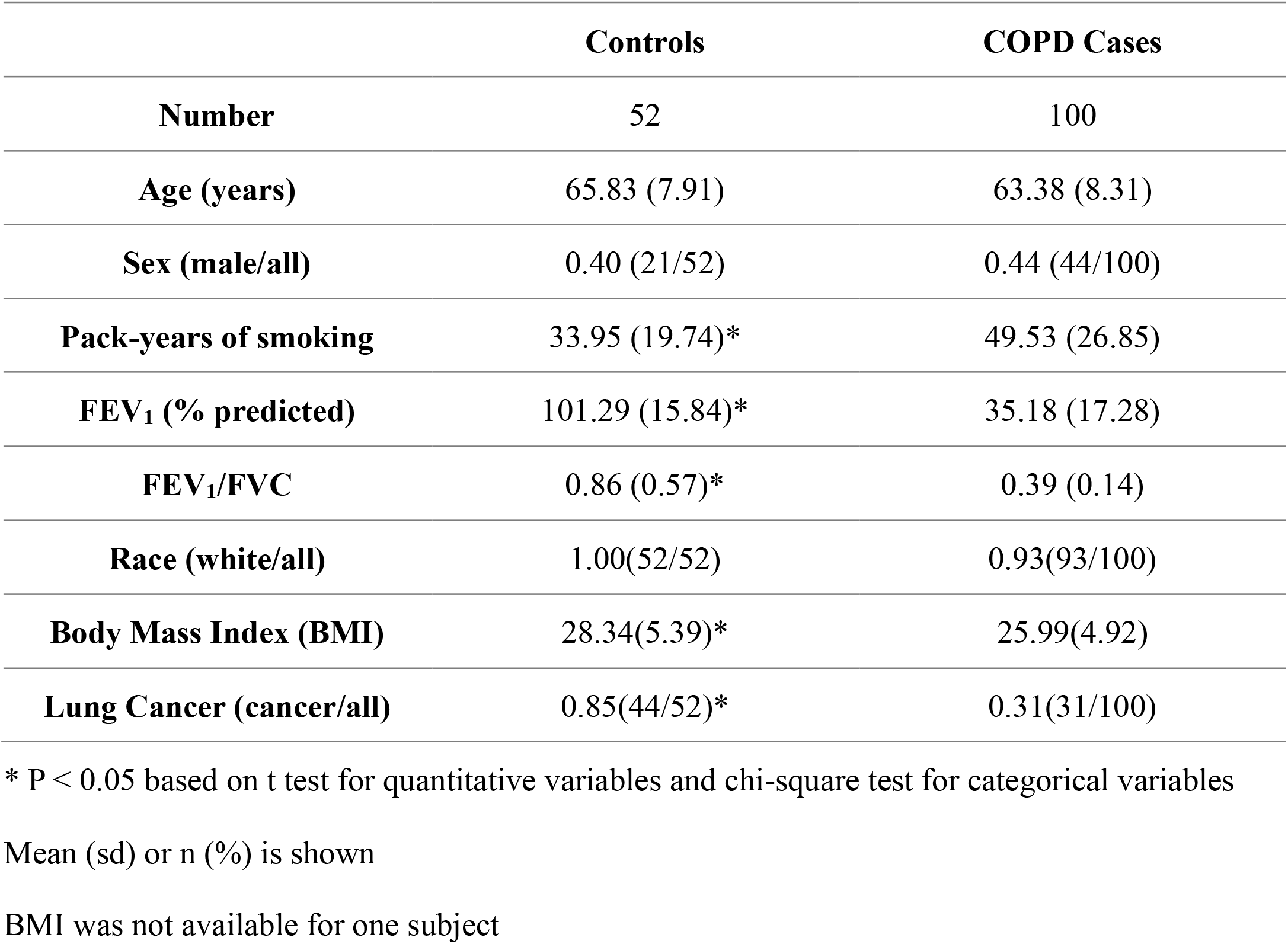
Clinical characteristics of study population.

|  | **Controls** | **COPD Cases** |
| --- | --- | --- |
| **Number** | 52 | 100 |
| **Age (years)** | 65.83 (7.91) | 63.38 (8.31) |
| **Sex (male/all)** | 0.40 (21/52) | 0.44 (44/100) |
| **Pack-years of smoking** | 33.95 (19.74)* | 49.53 (26.85) |
| **FEV<sub>1</sub> (% predicted)** | 101.29 (15.84)* | 35.18 (17.28) |
| **FEV<sub>1</sub>/FVC** | 0.86 (0.57)* | 0.39 (0.14) |
| **Race (white/all)** | 1.00(52/52) | 0.93(93/100) |
| **Body Mass Index (BMI)** | 28.34(5.39)* | 25.99(4.92) |
| **Lung Cancer (cancer/all)** | 0.85(44/52)* | 0.31(31/100) |

\* P < 0.05 based on t test for quantitative variables and chi-square test for categorical variables
Mean (sd) or n (%) is shown
BMI was not available for one subject

### Principal component analysis of proteomic data before and after preprocessing

We applied principal component analysis to assess the proteomic data distribution during preprocessing (filtering, removing outliers, normalization and imputation, and adding surrogate variables for batch effect correction). The effects of removing outliers and exogenous factors are shown in **Supplemental Figure S3**. After quality control, 4407 proteins in 150 samples were available for analysis.

### Linear regression analysis results (proteins with FDR < 0.05)

Linear regression models were established using the normalized and imputed datasets. We identified 25 proteins that differed significantly between COPD and control lung tissue samples with FDR < 0.05. The FDR values and beta-coefficients of these proteins are shown in **Table 2**. In addition, we included proteins associated at a less conservative FDR threshold (FDR < 0.1) in **Supplemental Table S9**. Among the top 25 proteins, most were expressed at lower levels in lung tissue from the subjects with COPD. For example, the protein expression levels of Agrin, Annexin A2, caveolin-1, and IL33 are negatively associated with COPD. Exceptions to this pattern were noted for the ferritin heavy and light chains, LDHA, and surfactant protein B, which were substantially higher in COPD cases. In the top 5% of proteins, there are 95 proteins positively and 125 proteins negatively associated with COPD. For comparison, we examined the linear regression results of several previously reported COPD protein biomarkers in **Table 3**. Trends for directionally consistent association (with p-value < 0.05 but FDR > 0.05) were noted for RAGE, fibrinogen, matrix metalloproteinase (MMP)-2, TGM2, and macrophage capping protein.

**Table 2:**
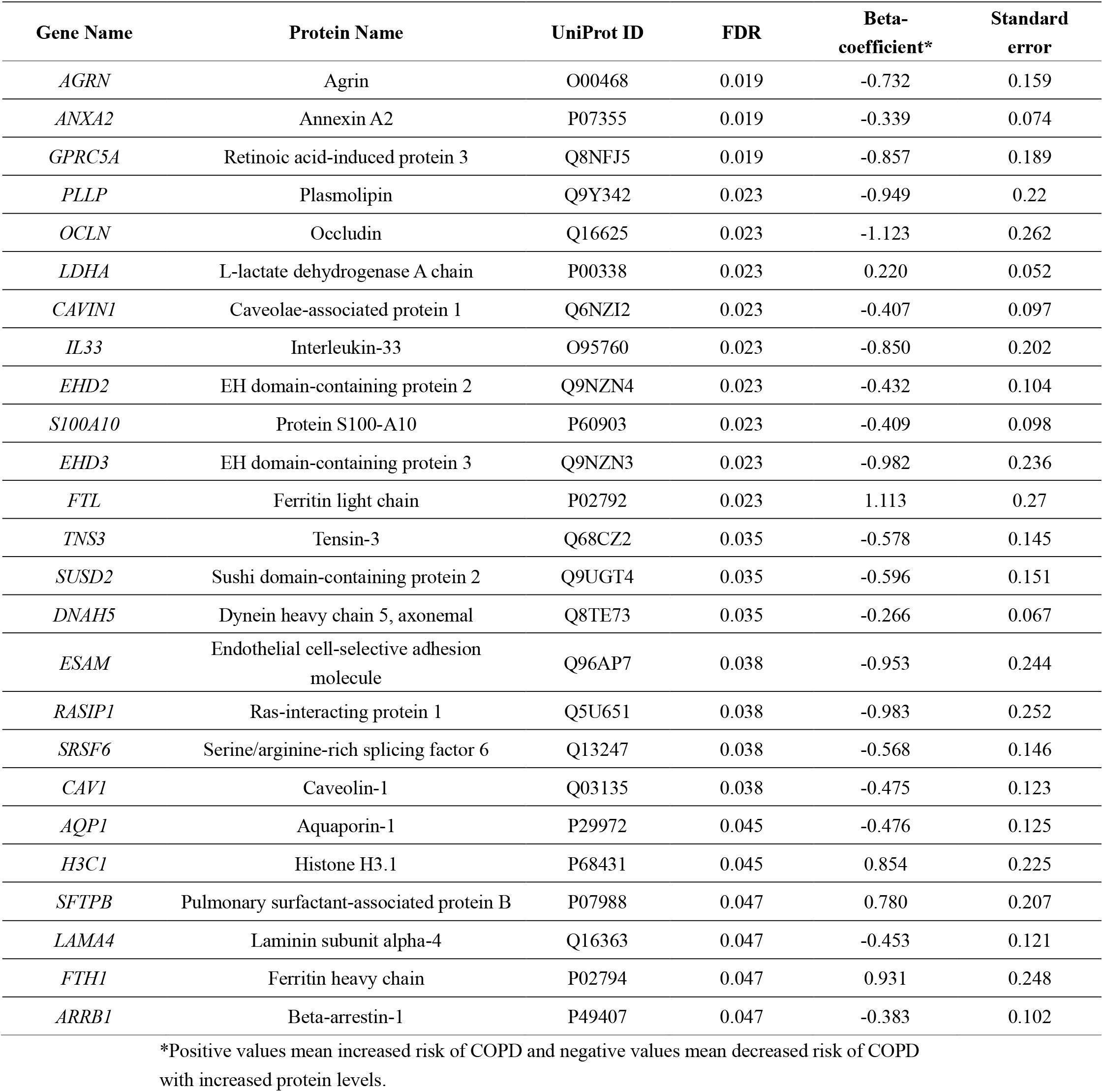
Top proteins associated with COPD based on linear regression analysis.

**Table 3:**
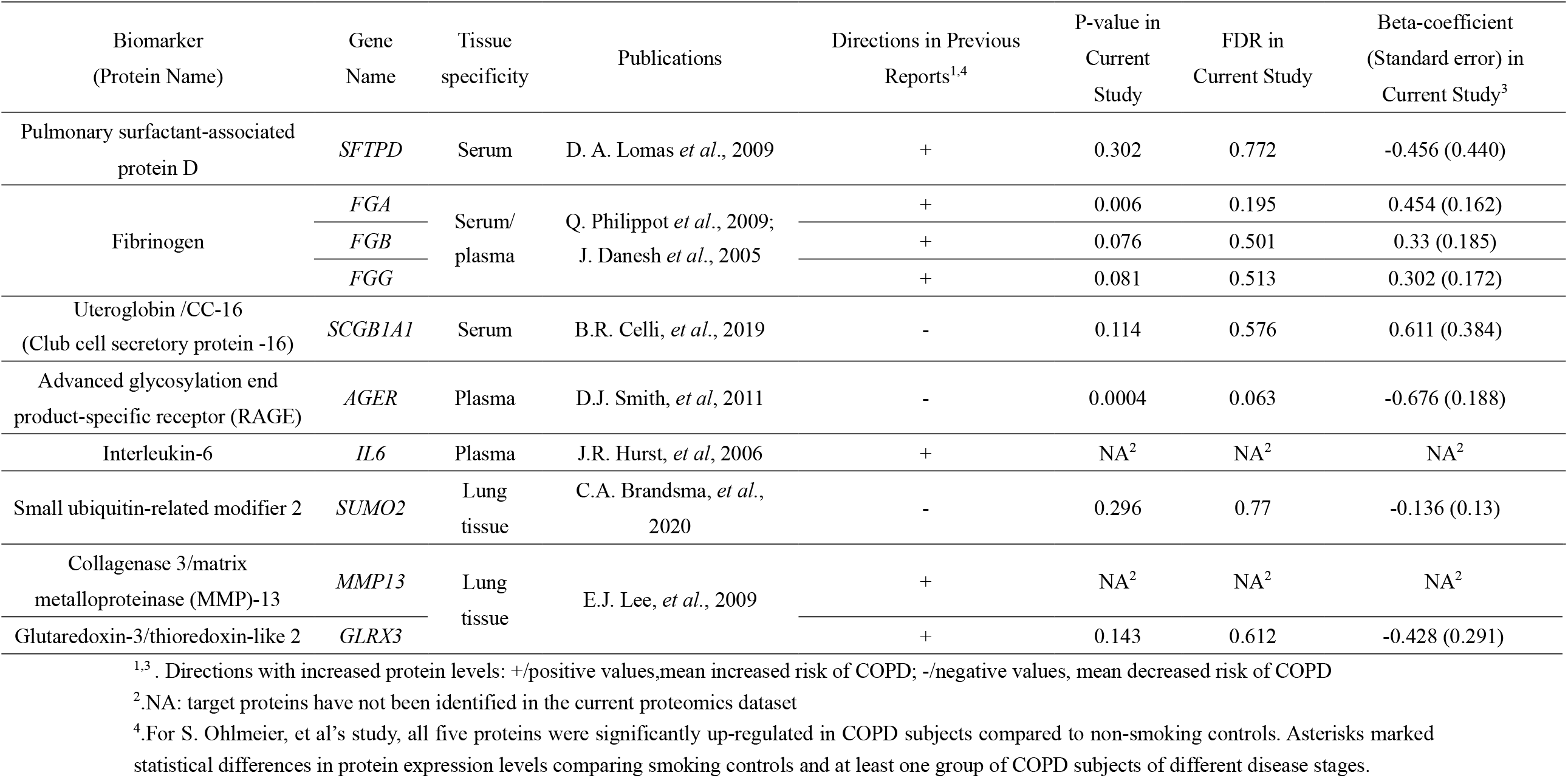
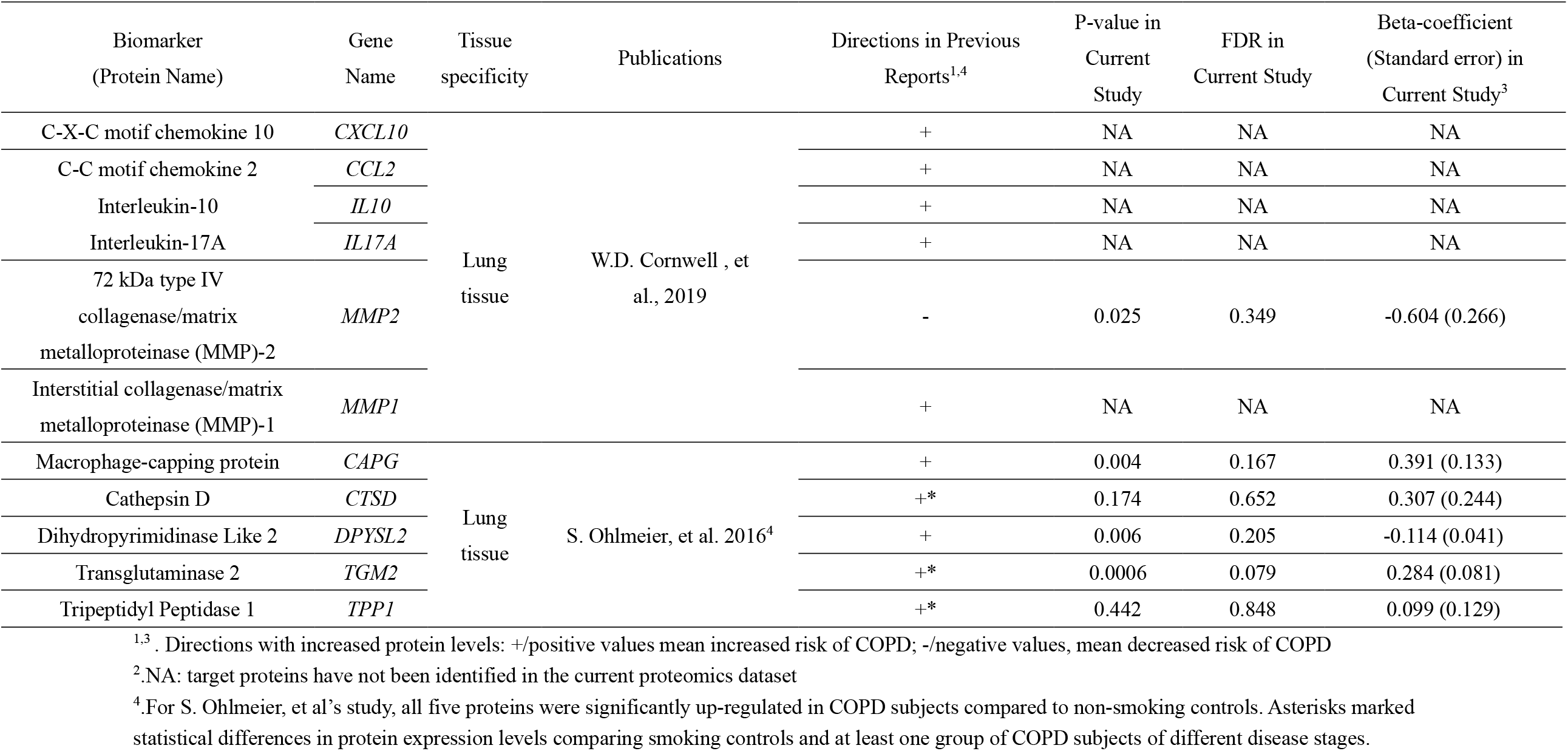
Linear regression results of selected previously reported COPD protein biomarkers.

### Correlations between top proteins from linear regression analysis

We calculated the pairwise Pearson correlation coefficients between the top 5% of COPD-associated proteins. The largest pairwise correlations are shown in **Table 4** (based on residuals after removing age, sex, batch effects, and surrogate variables). Also, we include a scatterplot to visualize the top correlations (**Supplemental Figure S4**) and a histogram of pairwise correlations (**Supplemental Figure S5**). Eleven high correlations (> 0.8) were noted. Some of the highest correlations involved proteins in shared molecular processes like FTL (Ferritin light chain) and FTH1 (Ferritin heavy chain), both part of ferritin, and CAV1 (Caveolin-1) and CAVIN1 (Caveolae-associated protein 1), both involved in caveolae formation. Additional correlation pairs like EHD2 (EH domain-containing protein 2) and RAGE (Advanced Glycosylation End-Product Specific Receptor), CAVIN1 and RAGE, and FTL and LGMN were also identified.

**Table 4:** Largest Pairwise Correlations between top 5% of proteins (after removing age, sex, batch effects and surrogate variables) associated with COPD.

| **Protein 1** | **Protein 2** | **Correlation**<br>**Coefficient (r)** | **P-value** |
| --- | --- | --- | --- |
| Ferritin light chain | Ferritin heavy chain | 0.921 | <0.001 |
| Caveolae-associated protein 1 | Caveolin-1 | 0.888 | <0.001 |
| EH domain-containing protein 2 | Caveolae-associated protein 1 | 0.881 | <0.001 |
| EH domain-containing protein 2 | Caveolin-1 | 0.866 | <0.001 |
| Caveolae-associated protein 1 | Advanced glycosylation end product-specific receptor | 0.864 | <0.001 |
| Complement C3 | Ceruloplasmin | 0.844 | <0.001 |
| Annexin A2 | Protein S100-A10 | 0.836 | <0.001 |
| EH domain-containing protein 2 | Advanced glycosylation end product-specific receptor | 0.834 | <0.001 |
| Caveolae-associated protein 1 | Caveolae-associated protein 2 | 0.821 | <0.001 |
| Ferritin light chain | Legumain | 0.820 | <0.001 |
| Caveolin-1 | Advanced glycosylation end product-specific receptor | 0.820 | <0.001 |

### Machine Learning Protein Biomarker Prediction of COPD

#### Identifying the best model through cross-validation

Our linear regression models only reflected the association between COPD and the expression level of one protein at a time. To study panels of proteins, we introduced machine learning models to establish multivariate prediction models for COPD. Three different machine learning methods were applied to develop predictive models distinguishing COPD subjects and controls: Random Forests, Naïve Bayes, and Elastic Net. Based on five-fold cross-validation, we calculated the accuracy and AUC to evaluate their performance. After comparison, the Random Forest method has the best performance based on both accuracy and AUC (**Table 5**).

**Table 5:**
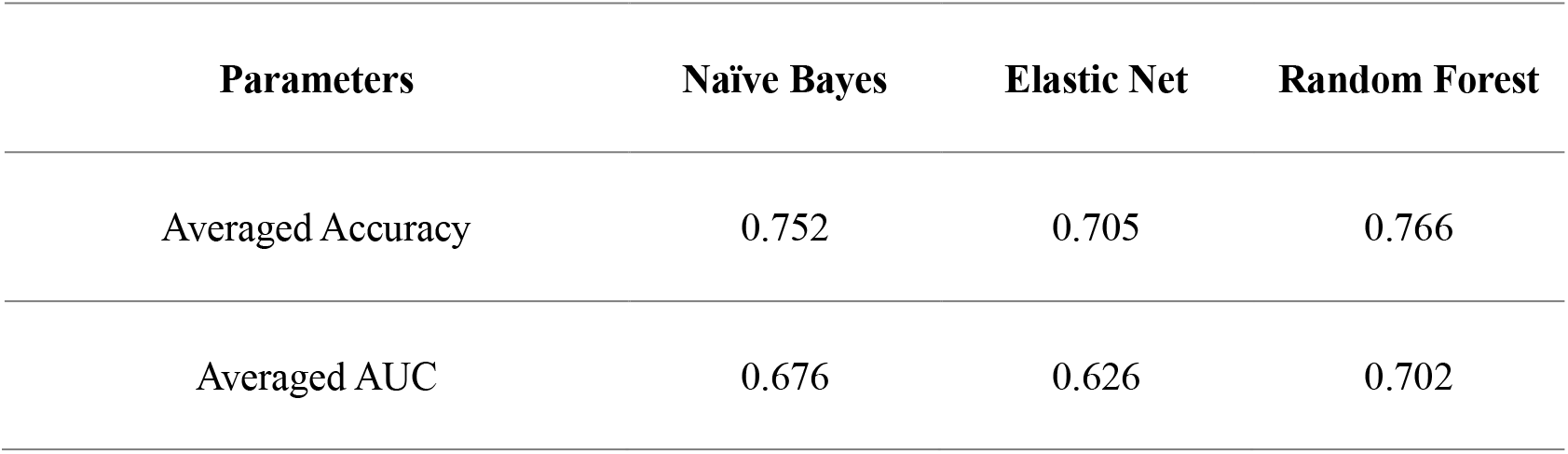
Performance and comparison of three machine learning methods for COPD prediction.

#### Feature importance evaluation

We evaluated the contribution of each Random Forest model feature to its COPD prediction performance. The feature importance of the top proteins and their original FDR in linear regression analysis are included in **Table 6**. Most proteins with high feature importance were also strongly associated in linear regression analyses. Some proteins with lower COPD association FDR also did achieve higher feature importance in Random Forests (e.g., BPIFA1, GPAA1).

**Table 6:**
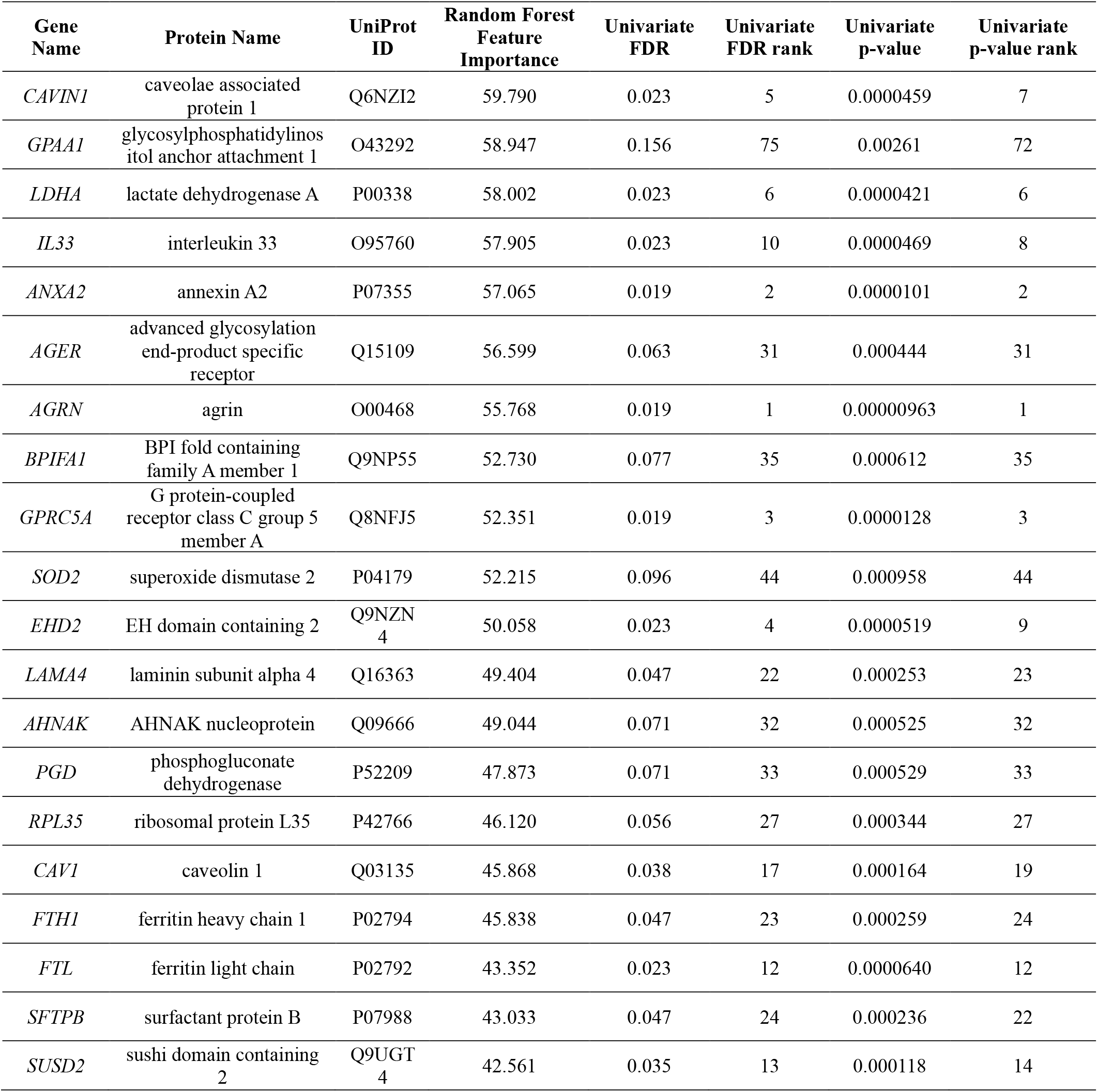
Feature importance evaluation of Random Forest model.

### Gene ontology enrichment analysis with candidate protein biomarkers for COPD

To investigate potential biological functions related to protein biomarkers, we chose the top 5% of proteins according to linear regression analysis for GO enrichment analysis (**Supplemental Table S10**). The enriched GO terms with p < 0.001 can be seen in **Figure 1**. Of interest, extracellular exosomes, phospholipase A2 inhibitor activity, and cell adhesion were three of the top processes, suggesting that protein secretion in exosomes, phospholipase A2 inhibition, and cell adhesion could be relevant for COPD pathogenesis.

**Figure 1:**
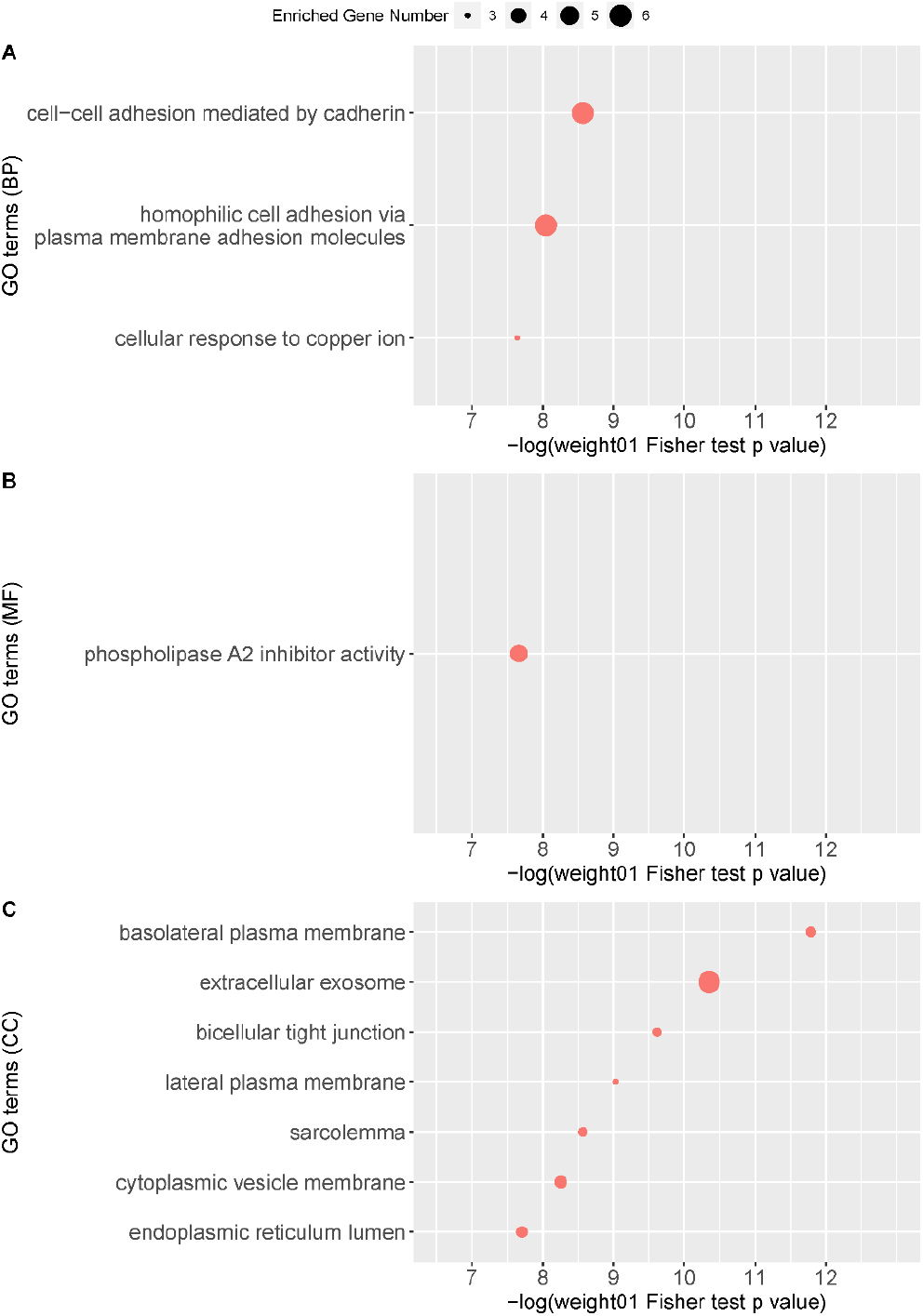
Gene ontology enrichment analysis on candidate protein biomarkers for COPD (top 5% of proteins according to FDR in linear regression analysis). GO enrichment analysis was performed on candidate protein biomarkers at three levels: A) biological processes (BP); B) molecular functions (MF); and C) cellular components (CC) with weight01 score threshold. The x-axis indicates the negative log value of weight01 Fisher test p-value. The more significantly the GO term enriched, the higher the value on the x-axis. The y-axis indicates different GO terms ranked by the weight01 Fisher test p-value. The size of the point in the figure indicates the number of genes significantly enriched in this GO term.

Additional analyses on the positively (n=95) and negatively (n=125) COPD-associated proteins in the top 5% of proteins were also performed. The enriched GO terms with p < 0.001 can be seen in **Supplemental Figure S6** and **Supplemental Figure S7** with detailed results listed in **Supplemental Table S11 and Supplemental Table S12**. Substantial differences were observed, with positively COPD-associated proteins enriched for processes including neutrophil degranulation and extracellular exosomes, while negatively COPD-associated proteins were enriched for processes including plasma membranes and cell-cell adhesion by cadherin.

## Discussion

Although identification of protein biomarkers for COPD and COPD-related phenotypes has been a topic of intensive investigation, few studies have utilized mass spectrometry for large-scale assessments in lung tissue samples. We found multiple protein biomarkers associated with COPD in lung tissue. However, these lung tissue biomarkers were largely distinct from previously reported plasma protein biomarkers. Our analysis had four parts, focusing on the top proteins associated with COPD, correlations between top proteins, performance of machine learning models, and functional enrichment analysis.

### Top proteins associated with COPD

We identified 25 protein biomarkers significantly associated with COPD. Other researchers have previously utilized mass spectrometry-based proteomics to investigate COPD in small sets of lung tissue samples. Matrix metalloproteinase (MMP)-13 and thioredoxin-like 2 (GLRX3) were found to be elevated in COPD with 22 subjects (E. J. Lee et al., 2009). We were unable to replicate the association to GLRX3, and we could not reliably detect MMP13. Cornwell and colleagues studied 34 proteins in lung tissue from subjects with combined pulmonary fibrosis and emphysema (CPFE; n=5), idiopathic pulmonary fibrosis (IPF; n=5), emphysema (n=5), and normal lungs (n=6) (Cornwell et al., 2019). Inflammatory factors like IL-6 and CCL2 were up-regulated in emphysema, whereas other factors like MMP2 were down-regulated (Cornwell et al., 2019); we found similar results for MMP2. Brandsma and colleagues integrated proteomics and transcriptomics analysis (Control, n = 8; COPD cases, n = 10) and identified SUMO2 as a candidate COPD protein biomarker (Brandsma et al., 2020). Their study identified 327 differentially expressed proteins in COPD lung tissue; these included EHD3, EHD2, ANXA2, TNS3, RASIP1, ARRB1, and S100A10 from our top 25 proteins—which had consistent directions of effect in our study. Five COPD-specific proteins (TGM2, CAPG, Cathepsin D, TPP1, and DPYSL2) were identified in small sets of lung tissue samples from several different diseases (smoking control, non-smoking control and IPF: n = 9 each, mild-moderate COPD, severe COPD, and alpha-1 antitrypsin deficiency: n = 8 each) by Ohlmeier and colleagues (Ohlmeier et al., 2016). We also identified TGM2 and CAPG with a consistent effect direction and nominally significant p-value < 0.05. Apart from studies focusing on COPD, several lung tissue studies have evaluated proteomic signals for COPD subtypes (such as alpha-1 antitrypsin deficiency) (Ohlmeier et al., 2016) and COPD-associated phenotypes (such as COPD exacerbations) (Sun, Ye, Wang, Bai, & Zhao, 2019). With the limited clinical phenotyping available in our study population, we were unable to address these questions.

Among the 25 proteins biomarkers in our analysis, several proteins have been previously implicated in COPD pathogenesis. IL33 is a cytokine serving as a mediator of chronic lung inflammation in COPD (Gabryelska, Kuna, Antczak, Białasiewicz, & Panek, 2019), but we found reduced IL33 levels in COPD lung samples. Agrin gene expression levels were found to be lower in lung tissue samples from severe COPD subjects (Xiao, Shu, Zhou, & Han, 2018), similar to our results. Plasma levels of the pro-peptide of surfactant protein B have been associated with COPD-related phenotypes (Leung et al., 2015). Previous studies (Fujimoto et al., 2012; Guo et al., 2019; Tao et al., 2007) confirmed that depletion of GPRC5A can promote inflammatory reactions in lung tissue. GPRC5A protein levels were found to be down-regulated in the lung tissue of COPD patients (Fujimoto et al., 2012), in agreement with our results. ANXA2 levels were found to be elevated in bronchoalveolar lavage (BAL) fluid in COPD subjects (Pastor et al., 2013), but we found lower levels in COPD lung tissue. Consistent with our results, Caveolin-1 has been reported to be down-regulated in COPD patients compared to normal controls in lung tissue samples (E. J. Lee et al., 2009). *DNAH5* mutations can cause primary ciliary dyskinesia, which can include bronchiectasis, and DNA variations in the region may also be associated with total lung capacity in COPD subjects (J. H. Lee et al., 2014). Further studies of these candidate protein biomarkers may assist in understanding COPD pathogenesis.

Interestingly, increased levels of two ferritin peptides (FTL and FTH1) encoding the same protein complex were associated with COPD in our dataset. Ferritin-associated proteins were previously reported to be up-regulated in the BAL fluid (Ghio et al., 2008) and alveolar macrophages (Philippot et al., 2014) of smoking COPD patients compared to healthy smokers, which is consistent with our results. Studies of one of the top COPD GWAS genes, *IREB2*, have focused on the role of iron-related pathways in COPD pathogenesis (Suzanne M Cloonan et al., 2016; S. M. Cloonan et al., 2017). Determining whether these findings are related to the same underlying regulatory mechanisms in COPD pathogenesis requires further study.

The most consistently associated plasma protein biomarker with emphysema, RAGE (Yonchuk et al., 2015), did not meet our FDR threshold for significance. However, RAGE was nominally significant (Beta = −0.68, P = 0.0004, FDR = 0.063), with lower expression levels in COPD lung tissue. Previous studies have confirmed negative correlations between the soluble isoform (sRAGE) and COPD (Cheng et al., 2013; Cockayne et al., 2012). Interestingly, the soluble and receptor forms of RAGE actually have opposite biological functions. Soluble RAGE plays a protective role against COPD pathogenesis (Yonchuk et al., 2015), blocking the classic RAGE signaling pathway. However, the receptor form of RAGE promotes chronic inflammatory diseases including COPD (Hofmann et al., 1999; Yonchuk et al., 2015). The mixed detection of soluble and receptor RAGE may affect the quantitative results of mass spectrometry. We were unable to distinguish sRAGE and RAGE in our peptide analysis. Similarly, we were unable to identify different forms of another previously reported COPD-associated protein, ICAM1(Zandvoort et al., 2006). Further quantitative detection of different forms of RAGE and ICAM1, such as through more sensitive MS approaches utilizing targeted assays (Kusebauch et al., 2016), may help clarify their significance.

We adjusted our association analyses of COPD case-control status in our study population of ex-smokers for age, sex, and technical factors. We did find confounding between COPD case-control status and pack-years (higher pack-years in COPD cases) and lung cancer diagnosis (more lung cancer in control subjects). Although some evidence for association to COPD of the top 25 proteins remained after adjusting for these confounders, the associations were often attenuated. Further investigation in other study populations will be required to determine if any of these top COPD-associated proteins are primarily related to lung cancer or smoking intensity.

### Correlations between top proteins

Among the top 5% of proteins associated with COPD, we identified 11 pairs of highly correlated proteins (correlation coefficient > 0.8). Some of these proteins were parts of the same protein complex or biological process, such as FTL and FTH1 forming the ferritin complex, and CAVIN1 and CAV1 involved in caveolae formation. Of the 11 pairs of highly correlated proteins, EHD2 and CAVIN1 were each highly correlated with three other proteins, and both of them were correlated with RAGE (Miniati et al., 2011; Sukkar et al., 2012). Highly correlated proteins could directly contribute to the pathogenesis of COPD by participating in similar COPD related pathways, and /or be co-expressed for reasons not due to COPD pathogenesis.

### Performance of machine learning models

To identify potential combinations of protein biomarkers related to COPD, we established three machine learning models using cross-validation. Considering both accuracy and AUC results, Random Forest provided the best performance in our proteomic dataset. In several recent publications, Random Forest has been applied in multiple mass spectrometric-based proteomics studies with relatively good performance (Izmirlian, 2004; Swanson, Xu, Nettleton, & Glatz, 2012). Apart from the prediction performance evaluation, we also obtained an importance ranking of features that contributed to COPD prediction in Random Forest models. Most of the top proteins in this list, including AGRN (rank 1 in FDR list and rank 7 in feature importance list), ANXA2 (rank 2 in FDR list and rank 5 in feature importance list), GPRC5A (rank 3 in FDR list and rank 9 in feature importance list) and IL33 (rank 8 in FDR list and rank 4 in feature importance list) were also top features in the linear regression analysis. Apart from these proteins with FDR < 0.05, the previously widely-reported COPD-associated gene *AGER* (RAGE) was also highlighted by the Random Forest model with rank 6. These results indicate that machine learning models verified the significance of many biomarkers identified in single protein analysis. Further investigation of proteins like GPAA1 (rank 2) and BPIFA1 (rank 8) that were highly important in Random Forest analysis but less significant in linear regression analysis may be warranted. Future studies of these machine learning models in plasma samples could lead to clinically relevant protein prediction models.

### Functional enrichment analysis

GO enrichment analyses were performed on the top 5% of proteins for functional exploration. Several biological processes associated with cell adhesion were found, including GO:0044331 (cell-cell adhesion mediated by cadherin) and GO: 0007156 (homophilic cell adhesion via plasma membrane adhesion molecules). According to the Gene Ontology Resource (The Gene Ontology Consortium, 2019), multiple cadherin-associated proteins (e.g., cadherin 1, cadherin 2, and cadherin 3) are included in these GO pathways. Cadherins are calcium-dependent proteins regulating cell adhesion processes (Meigs, Fedor-Chaiken, Kaplan, Brackenbury, & Casey, 2002). Further, cadherin has also been reported to participate in the epithelial to mesenchymal transition in smokers and subjects with COPD (Eapen et al., 2018).

Among GO molecular function terms, only one term, GO:0019834 (phospholipase A2 inhibitor activity), was identified. COPD-associated oxidative stress was reported to promote the activity of phospholipase A2 (Boukhenouna, Wilson, Bahmed, & Kosmider, 2018; Pniewska & Pawliczak, 2013). The inhibition of phosphodiesterase 4 signaling, involving phospholipase A2, can inhibit neutrophilic inflammation in COPD (Meliton et al., 2006).

Among GO terms for cellular components, GO:0070062 (extracellular exosomes) was identified. Extracellular exosomes are a type of extracellular vesicle released by multiple cell types. A systematic review of extracellular vesicles in the lung microenvironment noted that extracellular exosomes participate in the regulation of airway inflammation (Fujita, Kosaka, Araya, Kuwano, & Ochiya, 2015). Further, the inhibition of extracellular vesicles (including exosomes) was reported to mediate autophagy, which indirectly controls COPD-associated processes like airway remodeling (Fujita, Araya, et al., 2015). Of interest, the cellular processes related to the proteins that were positively and negatively associated with COPD were substantially different; further investigation of the pathobiological implications of these findings will be required.

### Limitations of the current study

Although our study has provided some progress in COPD proteomics analysis, we acknowledge several important limitations. We included lung tissue samples from 150 subjects. Although this is substantially larger than previous proteomic studies of lung tissue (Brandsma et al., 2020; Cornwell et al., 2019; E. J. Lee et al., 2009; Ohlmeier et al., 2016), our sample size may be inadequate for comprehensive identification of COPD protein biomarkers. Missing values were problematic, similar to other mass spectrometry proteomic studies. These missing values could relate to low abundance proteins near the lower limit of detection, variable quality of the lung tissue samples, or disease-related variability in protein levels. We used the same proteomics dataset to identify potential COPD biomarkers, build machine learning models, and evaluate model performance. The availability of replication datasets will be important to identify reliable biomarkers and establish stable prediction models. The lung tissue samples were obtained during thoracic surgical procedures for clinical indications that could influence proteomic profiles. Confounding of COPD case-control status with pack-years of smoking and lung cancer in our study population is a limitation of our studies. With limited clinical phenotyping and a modest sample size, we were unable to examine COPD subtypes or COPD-related phenotypes. In addition, we acknowledge that the subset of COPD patients undergoing thoracic surgery is likely not representative of the general COPD population. Finally, use of bulk lung tissue samples does not allow us to determine if our results were related to changes in cellular composition or to changes of protein levels within specific cell types.

In summary, despite a moderate sample size of lung tissue specimens (yet the largest cohort to be analyzed to date), we identified 25 potential protein biomarkers associated with COPD. Some of these biomarkers have been previously related to COPD pathogenesis. Several of the top proteins were highly correlated. Except for RAGE and fibrinogen, we did not find evidence that previously reported COPD plasma protein biomarkers were differentially expressed in COPD lung tissue. Future studies should involve larger sample sizes, replication populations, and assessments of identified lung tissue biomarkers in more readily available biospecimens (e.g., plasma).

## Data Availability

Data will be available through the Proteomics Identification Database at EMBL-EBI upon manuscript publication.

## Author contributions

Conception and design: EKS, RLM, YHZ

Analyses and interpretation: YHZ, MRH, PJC, KAS, MKM, MHC, JL, RLM, EKS

Drafting the manuscript for important intellectual content: All authors

## Acknowledgements

The study is supported by NIH grants, including R01 HL133135, R01 HL147148, R01 HL137927, R01 HL124233, R01 GM087221 and P01 HL114501.

This study utilized biological specimens and data provided by the Lung Tissue Research Consortium (LTRC) supported by the National Heart, Lung, and Blood Institute (NHLBI).

## Conflict of interest Statement

In the past three years, EKS, PJC, and MHC have received grant support from GSK and Bayer. MHC has received speaking or consulting fees from AstraZeneca and Illumina. PJC has received consulting fees from GSK and Novartis. Other authors do not have a financial relationship with a commercial entity that has an interest in the subject of this manuscript.

## Notes

### Author Declarations

the Partners Human Research Committee as the IRB with protocol number 2013P000706/BWH

