## Supplementary Materials for "Lung proteomic biomarkers associated with chronic obstructive pulmonary disease"

ONLINE DATA SUPPLEMENT

**Supplementary Materials**

**Sample Preparation**

Protein was extracted from the 152 lung tissue samples through mechanical shearing with a Precellys 24 homogenizer (Bertin instruments, France) using Ceramic 2.8 mm PowerBead Tubes (Qiagen Cat No. 13114-50), followed by cryolysis, in 50 mM Tris + 5% Sodium Dodecyl Sulfate (SDS) buffer. Protein quantity was measured using bicinchoninic acid assay (BCA assay), and 300 µg of protein from each sample was reduced with 5 mM Dithiothreitol, alkylated with 10 mM iodoacetamide, and digested with trypsin (Promega, USA, Gold Cat No. V5280), following SDS removal with an S-Trap Mini (Protifi, NY, USA). All chemicals are from Millipore-Sigma unless otherwise noted.

**Data Dependent Analysis Mass Spectrometry**

Each sample was analyzed in triplicate using Data Dependent Analysis (DDA) by high resolution nano LC-MS/MS (456 total LC-MS/MS runs). For each run, 1 µg of digested protein per sample was loaded onto a 2 cm Acclaim PepMap 100 trap column (75 um ID, C18 3 µm, (Thermo-Fisher Scientific USA)). The samples were analyzed using nano reverse-phase chromatography with a 50 cm fused silica column (PicoFrit 75 µm ID, New Objective) packed with ReproSil Pur 1.9 µm C18 (Dr. Maisch GmbH, Germany). Peptide elution was performed using an Easy nLC-1000 system (Thermo-Fisher Scientific). Mobile phase A consisted of 0.1% formic acid in water, and mobile phase B consisted of 0.1% formic acid in acetonitrile. A two-hour gradient elution was performed from 5% to 35% mobile phase B, followed by a 15 minute wash at 80% mobile phase B, and a 20 minute column re-equilibration at 5% mobile phase B. The flow rate was set at 300 nl/min and the column was heated at 60°C using a microtubing heater (MonoSLEEVE, Analytical Sales & Services Inc, USA). Mass spectra were acquired on a QExactive-HF (Thermo-Fisher Scientific, USA) mass spectrometer operated in top-15 DDA mode with dynamic exclusion. The precursor scan range was from 375-1375 *m/z* at 60,000 resolution and 3e6 target AGC with a 50 ms maximum injection time. A 1.8 *m/z* selection window was used to acquire MS/MS spectra at 15,000 resolution, an AGC target of 1e5, 100 ms maximum injection time, and fragmented using HCD with a normalized collision energy of 27. Dynamic exclusion was set to 20 seconds, with peptide match set to preferred and isotope exclusion turned on. Charge exclusion was set to 1 and greater than 5.

**Mass Spectrometry Data Analysis**

The raw mass spectra files were converted to mzML format using MSConvert from Proteowizard (Chambers et al., 2012) and analyzed using the Trans-Proteomic Pipeline (Deutsch et al., 2015). The analysis pipeline consisted of database searching with Comet (version 2016.01 rev. 2) (Eng, Jahan, & Hoopmann, 2013) against the *homo sapiens* UniProt (Apweiler et al., 2004) reviewed proteome (downloaded March 11, 2016) and shuffled decoy sequences. Comet parameters included a fixed modification of +57.021464 Da on cysteine and a variable modification of +15.994915 Da on methionine. A precursor tolerance of 25 ppm was set, with a fragment bin tolerance of 0.2 and fragment bin offset of 0. Complete enzymatic cleavage with up to 2 missed cleavages was set. Peptide-spectrum matches (PSMs) were validated using PeptideProphet (Keller, Nesvizhskii, Kolker, & Aebersold, 2002) and iProphet (Shteynberg et al., 2011). Peptide precursor ion quantitation was performed following a 1% false discovery rate threshold across all 456 sample runs. The area under the chromatogram for each precursor ion was extracted using an in-house mzML spectra parsing algorithm. Additionally, retention time alignment and mass matching was used to extract precursor ion signals using identifications from adjacent runs, similar to other approaches (Cox et al., 2014). Precursor ion abundances between runs were quartile normalized, and summed together when observed over multiple charge states. Protein quantity was then estimated as the sum of up to the top three most abundant peptides among the proteotypic peptides (i.e., peptides that match to only a single protein), using the same peptides for each protein in each sample.

**Proteomics Data analysis**

***Comprehensive workflow for proteomics datasets***

We summarized the entire comprehensive workflow for the proteomics datasets integrating data preprocessing, linear regression analysis, machine learning analysis, correlation analysis and gene ontology enrichment analysis in **Supplemental Figure S1**.

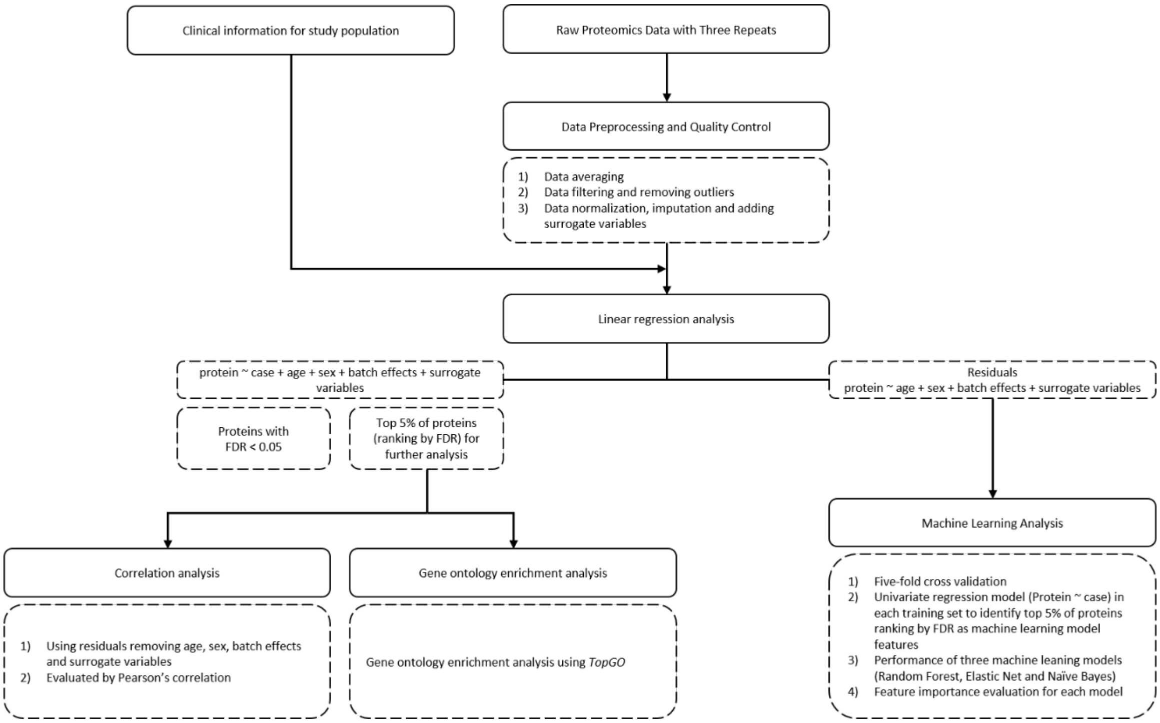


**Figure S1: Workflow for proteomics analysis.** Description of the workflow for proteomics dataset analyses. We performed data preprocessing, data quality control and linear regression analysis to identify the top COPD associated proteins. Using the top 5% of associated proteins (n=220), correlation analysis, machine learning model-based multivariate analysis, and functional gene ontology enrichment analysis are performed to identify co-expressed COPD-associated proteins, panels of predictive proteins, and over-represented biological pathways in COPD-associated proteins.

***Data preprocessing on raw proteomics datasets***

**1) Data averaging:** The raw proteomics datasets with three repeated measurements for each lung tissue sample were averaged to obtain a final value for each protein in each sample. For proteins with missing values, we only averaged the available values (removing missing values).

**2) Data filtering:** Then, we calculated the proportion of missing values (zeros) in raw proteomics datasets for each protein and removed proteins missing in more than 50% of samples (**Supplemental Figure S2**). Further, based on the distribution of missing values among samples, we identified and removed two outlier lung tissue samples from our original datasets following **the interquartile rule for**

**outliers (Han, Pei, & Kamber, 2011):**

① Lower outlier = Q1 – (1.5 × IQR)；

② Higher outlier = Q3 + (1.5 × IQR) (Q1, Q3 the first and third quartiles, IQR = Q3 – Q1).

**3) Data normalization, imputation, and surrogate variables:** After removing proteins with many missing values (>50%) and outlier samples, the R package *vsn* (Variance Stabilization and Calibration for Microarray Data) (version 3.56.0) (Huber, Von Heydebreck, Sültmann, Poustka, & Vingron, 2002) was used for the normalization of filtered datasets, following previous reports on the normalization of quantitative label-free proteomics datasets (Välikangas, Suomi, & Elo, 2018). After normalization, the R package *vim* (Visualization and Imputation of Missing Values) (version 6.0.0) (Torgo, 2016) was applied to perform K-Nearest Neighbor (KNN) imputation on the normalized dataset. Next, we applied the R package *sva* (Surrogate Variable Analysis) (version 3.36.0) (Leek, Johnson, Parker, Jaffe, & Storey, 2012) to create the surrogate variables while specifying covariates of age, sex, and columns (batch effects). Two surrogate variables (sv1 and sv2) were generated for the correction of datasets after imputation and normalization. The distribution of missing values before and after filtering are in **Supplemental Figure S2**. Plots were created with *ggplot2* (version 3.3.2) R packages.

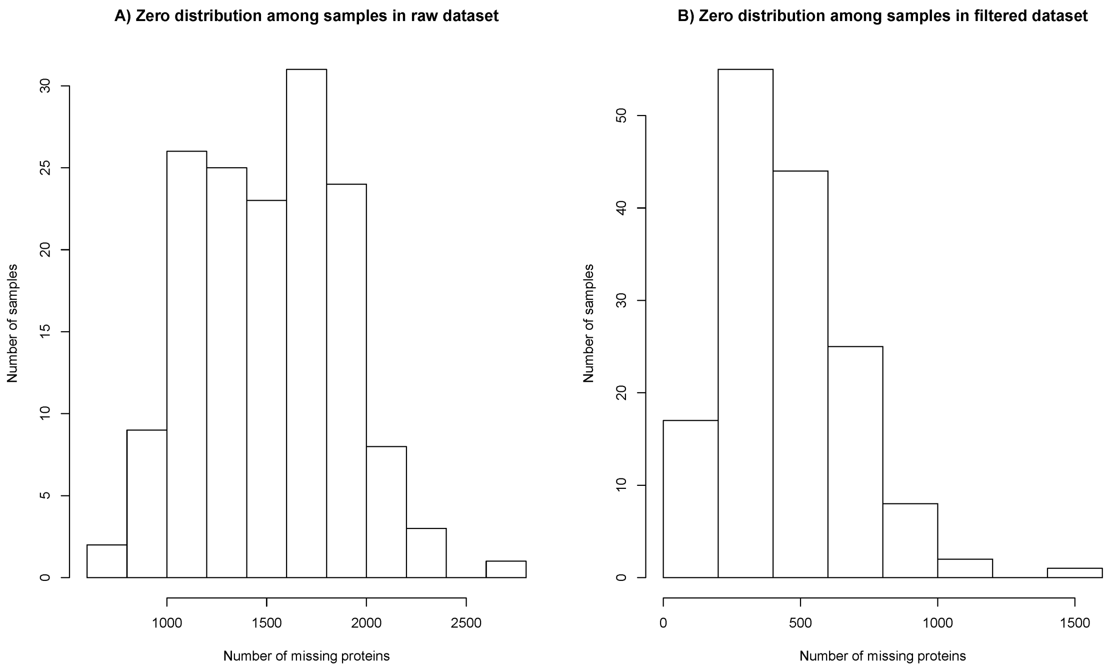


**Figure S2: Number of zero values in raw and filtered proteomics datasets.** A) Number of zero values among different samples in raw datasets; B) Number of zero values among different samples after removing proteins with more than 50% missing data.

***Principal component analysis for data before and after preprocessing***

We applied principal component analysis to assess the distribution pattern of our proteomics datasets during preprocessing (data filtering, outlier removing, normalization, imputation, and adding surrogate variables). The detailed data distribution patterns during the preprocessing procedure are shown in **Supplemental Figure S3**. Plots were created with the *ggplot2* (version 3.3.2) R package.

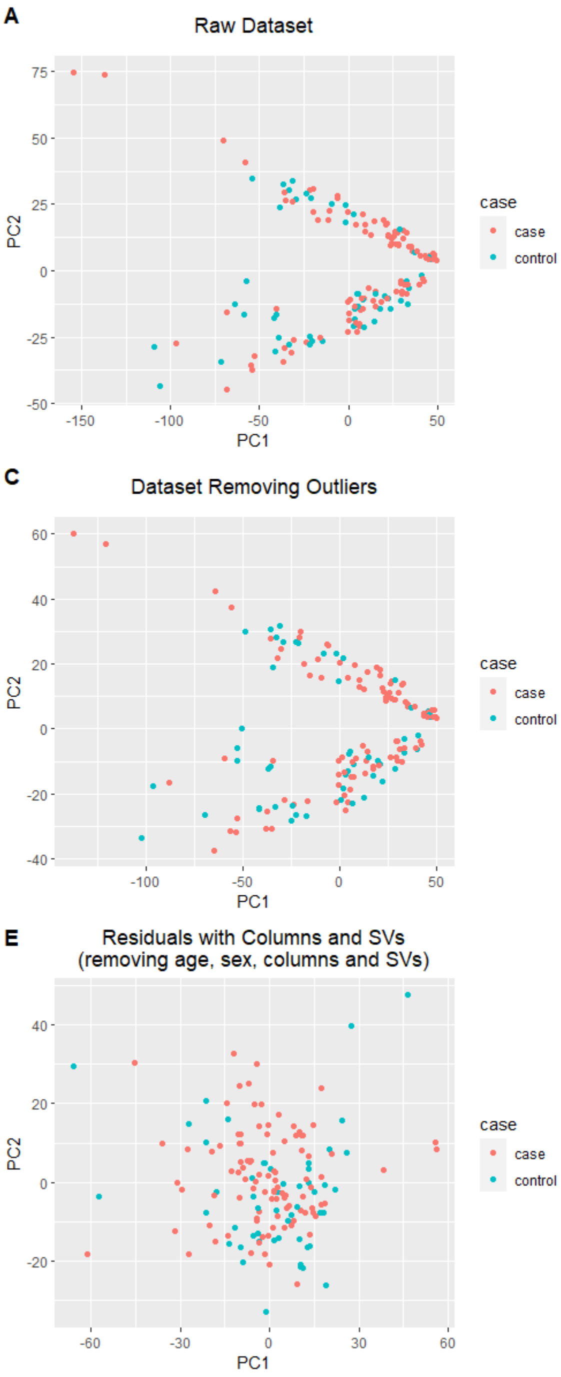


**Figure S3: Principal Component Analysis (PCA) plots of the data distribution during preprocessing.** A) raw dataset; B) raw dataset removing proteins with more than 50% missing values; C) dataset removing two outlier samples with large amounts of missing data; D) residuals removing effects of sex, age, and column number (batch effects); E) residuals removing effects of sex, age, column number (batch effects) and surrogate variables.

***Finding optimal covariates for linear regression analyses of COPD protein biomarkers***

We examined potential covariates of age, sex, pack-years of smoking, race, BMI, and lung cancer status for proteomic analysis of COPD. To find optimal covariates for the linear regression models, we evaluated the importance of each covariate by several methods.

Firstly, according to Table 1, only 7 subjects of 100 COPD cases are non-White and no subjects in the control group are non-White, as shown in **Supplemental Table S1**. Therefore, we elected not to include race as a covariate in our COPD proteomic analysis.

Secondly, to evaluate the remaining candidate covariates, we built a series of linear regression models using one covariate at a time and summarized the p-values of COPD case/control status and the covariates and the beta-coefficients of COPD case/control status to evaluate the contribution of each covariate on the prediction of COPD-associated protein expression levels. These analyses were performed for the top 25 proteins associated with COPD selected from a model with age, sex, batch effects, and surrogate variables. Based on five covariates (age, sex, BMI, pack-years of smoking, and lung cancer), we established five linear regression models as shown below:

$L1\colon protein \sim case\ (0/1) + age + column(batch\ effects) + surrogate\ variables\ (SVs)$

$L2\colon protein \sim case\ (0/1) + sex + column(batch\ effects) + surrogate\ variables\ (SVs)$

$L3\colon protein \sim case\ (0/1) + BMI + column(batch\ effects) + surrogate\ variables\ (SVs)$

$L4\colon protein \sim case\ (0/1) + packyears + column(batch\ effects) + surrogate\ variables\ (SVs)$

$L5\colon protein \sim case\ (0/1) + cancer + column(batch\ effects) + surrogate\ variables\ (SVs)$

The results for COPD affection status p-value, beta-coefficients (standard error), and covariate p-value are shown in **Supplemental Table S3, S4 and S5**. The column names indicate the included covariates in the regression models. An additional column indicating the results of the original linear regression model (with age, sex, batch effects, and surrogate variables as covariates) was also added for comparison (L0). For these 25 proteins, adding lung cancer status as a covariate for the linear regression model (L5) often strongly decreased the significance of COPD case/control status (p-value), but all of these proteins remain at least nominally associated with COPD ($p<0.05$). As shown in **Supplemental Table S2**, lung cancer is confounded with COPD affection status. While COPD subjects often underwent thoracic surgery for reasons other than lung cancer (e.g., lung transplant, lung volume reduction surgery), control subjects usually were found to have lung cancer at thoracic surgery. The p-value of the covariate for lung cancer is quite significant for some proteins like Agrin and Plasmolipin. Both Agrin (Chakraborty et al., 2015) and Plasmolipin (Bach et al., 2018) have been suggested to be involved in tumorigenesis. To a lesser extent, we also found that including pack-years of smoking attenuated the association evidence for COPD, although all of these proteins remained associated with COPD at $p<0.01$. Pack-years of smoking was also confounded with COPD affection status, with COPD subjects having substantially higher mean pack-years of smoking than control subjects. Adding BMI, sex, or age as covariates did not substantially affect the P-values or beta coefficients of COPD case/control status with the top 25 protein levels.

Thirdly, we further evaluated the contribution of different candidate covariates on the protein levels of the top 25 proteins by examining the association of these covariates directly with protein levels without including COPD case/control status in the models. Linear regression models (L1',L2',L3',L4' and L5' ) were created as shown below:

$$L1': protein \sim age + column(batch\ effects) + surrogate\ variables\ (SVs)$$

$$L2': protein \sim sex + column(batch\ effects) + surrogate\ variables\ (SVs)$$

$$L3': protein \sim BMI + column(batch\ effects) + surrogate\ variables\ (SVs)$$

$$L4': protein \sim packyears + column(batch\ effects) + surrogate\ variables\ (SVs)$$

$$L5': protein \sim cancer + column(batch\ effects) + surrogate\ variables\ (SVs)$$

The p-values of each linear regression model reflect whether the specified covariate is associated with the protein expression level. The column rank presents the order of protein's p-value in the original linear regression model (with adjustment for age, sex, batch effects, and surrogate variables). These results are shown in **Supplemental Table S6**. The column names indicate the included covariates. For most top proteins (23/25), the p-value of lung cancer is nominally significant. As we have discussed above, such effects are related to confounding between COPD and lung cancer in our data set. These proteins might be associated with both COPD and lung cancer, or only with COPD or lung cancer. Many of the top proteins were at least nominally associated with pack-years of smoking as well.

To investigate further the effects of the confounding effects of lung cancer and pack-years of smoking, we also performed stratified analyses separately in COPD cases only and control subjects only. Although these smaller samples will likely have reduced power to detect associations, we investigated whether associations of lung cancer or pack-years of smoking to protein levels could still be detected. If the primary associations were with one of these covariates rather than with COPD, we expected to still see association within both of these strata. Thus, we established L4' and L5' again with the top 25 proteins in COPD cases or controls only and compared the P-values and beta coefficients of covariates as shown in **Supplemental Table S7 and S8.** The p-values for association of pack-years of smoking and lung cancer with protein levels are often nominally significant in either cases or controls. However, the p-values were not found to be consistently significant in both cases and controls. These results suggest that the primary associations of these proteins are with COPD rather than pack-years of smoking or lung cancer. Based on these analyses, we elected to include age and sex, as well as batch effects and surrogate variables, in our primary linear regression analyses.

**Table S1: COPD case/control and race**

| | Controls | COPD cases |
|---|---|---|
| White | 52 | 93 |
| Non-white | 0 | 7 |

*P-value for Fisher's exact test: 0.10

**Table S2: COPD case/control and lung cancer status**

| | Controls | COPD cases |
|---|---|---|
| With lung cancer | 44 | 31 |
| Without lung cancer | 8 | 69 |

*P-value for Fisher's exact test: $2.0 * 10^{-10}$

**Table S3: P-values for COPD of top proteins associated with COPD in linear regression model with COPD and one covariate**

**(Protein ~ COPD + covariate + batch effects + SVs)**

| Gene Name | Protein Name | Uniprot ID | Age | Sex | BMI | Pack-years | Cancer | Original |
|---|---|---|---|---|---|---|---|---|
| *AGRN* | Agrin | O00468 | 8.44E-06 | 3.07E-06 | 2.69E-06 | 2.94E-06 | 1.26E-02 | 9.63E-06 |
| *ANXA2* | Annexin A2 | P07355 | 9.35E-06 | 3.95E-06 | 7.14E-06 | 1.07E-04 | 1.34E-03 | 1.01E-05 |
| *GPRC5A* | Retinoic acid-induced protein 3 | Q8NFJ5 | 1.07E-05 | 1.74E-05 | 1.61E-05 | 6.96E-05 | 3.84E-03 | 1.28E-05 |
| *PLLP* | Plasmolipin | Q9Y342 | 3.30E-05 | 1.54E-05 | 4.12E-05 | 8.90E-05 | 8.47E-03 | 3.14E-05 |
| *OCLN* | Occludin | Q16625 | 2.97E-05 | 4.78E-05 | 4.94E-05 | 3.79E-04 | 1.54E-03 | 3.38E-05 |
| *LDHA* | L-lactate dehydrogenase A chain | P00338 | 3.68E-05 | 1.69E-05 | 1.78E-05 | 4.23E-05 | 1.58E-04 | 4.21E-05 |
| *CAVIN1* | Caveolae-associated protein 1 | Q6NZI2 | 4.26E-05 | 4.07E-05 | 2.45E-05 | 4.24E-04 | 3.24E-03 | 4.59E-05 |
| *IL33* | Interleukin-33 | O95760 | 4.17E-05 | 5.53E-05 | 4.14E-06 | 6.87E-04 | 1.07E-03 | 4.69E-05 |
| *EHD2* | EH domain-containing protein 2 | Q9NZN4 | 4.43E-05 | 4.04E-05 | 2.46E-05 | 4.91E-04 | 3.60E-03 | 5.19E-05 |
| *S100A10* | Protein S100-A10 | P60903 | 4.69E-05 | 2.90E-05 | 2.75E-05 | 7.90E-04 | 8.29E-04 | 5.43E-05 |
| *EHD3* | EH domain-containing protein 3 | Q9NZN3 | 5.04E-05 | 5.40E-05 | 9.73E-06 | 1.53E-04 | 2.49E-04 | 5.72E-05 |
| *FTL* | Ferritin light chain | P02792 | 6.10E-05 | 8.00E-05 | 5.94E-05 | 1.40E-03 | 7.39E-05 | 6.40E-05 |
| *TNS3* | Tensin-3 | Q68CZ2 | 8.90E-05 | 3.19E-04 | 1.34E-03 | 3.28E-04 | 4.61E-03 | 1.08E-04 |

**Table S3: P-values for COPD of top proteins associated with COPD in linear regression model with COPD and one covariate**

**(Protein ~ COPD + covariate + batch effects + SVs)**

**(Continued)**

| Gene Name | Protein Name | Uniprot ID | Age | Sex | BMI | Pack-years | Cancer | Original |
|---|---|---|---|---|---|---|---|---|
| *SUSD2* | Sushi domain-containing protein 2 | Q9UGT4 | 1.00E-04 | 6.23E-05 | 1.95E-05 | 5.49E-04 | 3.48E-02 | 1.18E-04 |
| *DNAH5* | Dynein heavy chain 5, axonemal | Q8TE73 | 1.03E-04 | 1.10E-04 | 1.70E-04 | 9.26E-04 | 4.90E-03 | 1.20E-04 |
| *ESAM* | Endothelial cell-selective adhesion molecule | Q96AP7 | 1.27E-04 | 6.67E-05 | 3.27E-05 | 1.51E-04 | 5.27E-04 | 1.46E-04 |
| *RASIP1* | Ras-interacting protein 1 | Q5U651 | 1.33E-04 | 8.80E-05 | 4.04E-05 | 3.92E-04 | 9.34E-04 | 1.50E-04 |
| *SRSF6* | Serine/arginine-rich splicing factor 6 | Q13247 | 1.33E-04 | 1.83E-04 | 1.09E-04 | 1.06E-03 | 1.43E-02 | 1.64E-04 |
| *CAV1* | Caveolin-1 | Q03135 | 1.45E-04 | 8.57E-05 | 6.31E-05 | 7.32E-04 | 1.05E-02 | 1.64E-04 |
| *AQP1* | Aquaporin-1 | P29972 | 1.95E-04 | 4.92E-05 | 2.49E-05 | 7.22E-04 | 3.49E-03 | 2.15E-04 |
| *H3C1* | Histone H3.1 | P68431 | 1.91E-04 | 1.49E-04 | 1.24E-04 | 1.29E-04 | 2.04E-03 | 2.16E-04 |
| *SFTPB* | Pulmonary surfactant-associated protein B | P07988 | 2.48E-04 | 1.77E-04 | 2.61E-04 | 1.36E-03 | 2.20E-03 | 2.36E-04 |
| *LAMA4* | Laminin subunit alpha-4 | Q16363 | 2.92E-04 | 1.27E-04 | 2.62E-04 | 3.41E-04 | 2.10E-02 | 2.53E-04 |
| *FTH1* | Ferritin heavy chain | P02794 | 2.46E-04 | 2.60E-04 | 1.13E-04 | 4.29E-03 | 4.56E-04 | 2.59E-04 |
| *ARRB1* | Beta-arrestin-1 | P49407 | 2.29E-04 | 5.76E-04 | 2.71E-04 | 1.56E-03 | 7.72E-03 | 2.67E-04 |

**Table S4: Beta-coefficients (standard error) for COPD of top proteins associated with COPD in linear regression model with COPD and one covariate**

**(Protein ~ COPD + covariate + batch effects + SVs)**

| Gene Name | Protein Name | Uniprot ID | Age | Sex | BMI | Pack-years | Cancer | Original |
|---|---|---|---|---|---|---|---|---|
| *AGRN* | Agrin | O00468 | -0.734 (0.159) | -0.764 (0.157) | -0.784 (0.16) | -0.8 (0.164) | -0.439 (0.174) | -0.732 (0.159) |
| *ANXA2* | Annexin A2 | P07355 | -0.339 (0.074) | -0.35 (0.073) | -0.349 (0.075) | -0.299 (0.075) | -0.272 (0.083) | -0.339 (0.074) |
| *GPRC5A* | Retinoic acid-induced protein 3 | Q8NFJ5 | -0.864 (0.189) | -0.829 (0.186) | -0.856 (0.192) | -0.802 (0.196) | -0.628 (0.213) | -0.857 (0.189) |
| *PLLP* | Plasmolipin | Q9Y342 | -0.943 (0.22) | -0.97 (0.217) | -0.943 (0.223) | -0.915 (0.227) | -0.656 (0.246) | -0.949 (0.22) |
| *OCLN* | Occludin | Q16625 | -1.126 (0.261) | -1.083 (0.258) | -1.108 (0.265) | -0.978 (0.268) | -0.963 (0.298) | -1.123 (0.262) |
| *LDHA* | L-lactate dehydrogenase A chain | P00338 | 0.225 (0.053) | 0.228 (0.051) | 0.237 (0.053) | 0.23 (0.054) | 0.234 (0.06) | 0.22 (0.052) |
| *CAVIN1* | Caveolae-associated protein 1 | Q6NZI2 | -0.407 (0.096) | -0.403 (0.095) | -0.424 (0.097) | -0.357 (0.099) | -0.326 (0.109) | -0.407 (0.097) |
| *IL33* | Interleukin-33 | O95760 | -0.852 (0.201) | -0.827 (0.199) | -0.941 (0.196) | -0.714 (0.206) | -0.768 (0.23) | -0.85 (0.202) |
| *EHD2* | EH domain-containing protein 2 | Q9NZN4 | -0.436 (0.103) | -0.431 (0.102) | -0.455 (0.104) | -0.377 (0.106) | -0.346 (0.117) | -0.432 (0.104) |
| *S100A10* | Protein S100-A10 | P60903 | -0.412 (0.098) | -0.417 (0.097) | -0.428 (0.099) | -0.339 (0.099) | -0.382 (0.112) | -0.409 (0.098) |
| *EHD3* | EH domain-containing protein 3 | Q9NZN3 | -0.986 (0.236) | -0.968 (0.232) | -1.073 (0.234) | -0.948 (0.243) | -1.011 (0.269) | -0.982 (0.236) |
| *FTL* | Ferritin light chain | P02792 | 1.111 (0.269) | 1.078 (0.265) | 1.126 (0.272) | 0.89 (0.273) | 1.25 (0.306) | 1.113 (0.27) |
| *TNS3* | Tensin-3 | Q68CZ2 | -0.589 (0.146) | -0.532 (0.144) | -0.479 (0.146) | -0.563 (0.153) | -0.483 (0.168) | -0.578 (0.145) |

**Table S4: Beta-coefficients (standard error) for COPD of top proteins associated with COPD in linear regression model with COPD and one covariate**

**(Protein ~ COPD + covariate + batch effects + SVs)**

**(Continued)**

| Gene Name | Protein Name | Uniprot ID | Age | Sex | BMI | Pack-years | Cancer | Original |
|---|---|---|---|---|---|---|---|---|
| *SUSD2* | Sushi domain-containing protein 2 | Q9UGT4 | -0.604 (0.151) | -0.611 (0.148) | -0.67 (0.152) | -0.547(0.155) | -0.355 (0.166) | -0.596 (0.151) |
| *DNAH5* | Dynein heavy chain 5, axonemal | Q8TE73 | -0.27 (0.067) | -0.263 (0.066) | -0.263 (0.068) | -0.234 (0.069) | -0.219 (0.077) | -0.266 (0.067) |
| *ESAM* | Endothelial cell-selective adhesion molecule | Q96AP7 | -0.958 (0.243) | -0.987 (0.24) | -1.053 (0.245) | -0.98 (0.252) | -0.985 (0.277) | -0.953 (0.244) |
| *RASIP1* | Ras-interacting protein 1 | Q5U651 | -0.987 (0.251) | -1.001 (0.248) | -1.055 (0.249) | -0.941 (0.259) | -0.97 (0.287) | -0.983 (0.252) |
| *SRSF6* | Serine/arginine-rich splicing factor 6 | Q13247 | -0.578 (0.147) | -0.555 (0.144) | -0.593 (0.149) | -0.508 (0.152) | -0.411 (0.166) | -0.568 (0.146) |
| *CAV1* | Caveolin-1 | Q03135 | -0.478 (0.122) | -0.488 (0.121) | -0.511 (0.124) | -0.434 (0.126) | -0.358 (0.138) | -0.475 (0.123) |
| *AQP1* | Aquaporin-1 | P29972 | -0.478 (0.125) | -0.525 (0.125) | -0.559 (0.128) | -0.448 (0.13) | -0.428 (0.144) | -0.476 (0.125) |
| *H3C1* | Histone H3.1 | P68431 | 0.859 (0.224) | 0.86 (0.221) | 0.894 (0.226) | 0.909 (0.231) | 0.805 (0.256) | 0.854 (0.225) |
| *SFTPB* | Pulmonary surfactant-associated protein B | P07988 | 0.775 (0.206) | 0.782 (0.203) | 0.781 (0.208) | 0.693 (0.212) | 0.729 (0.234) | 0.78 (0.207) |
| *LAMA4* | Laminin subunit alpha-4 | Q16363 | -0.448 (0.12) | -0.468 (0.119) | -0.458 (0.122) | -0.458 (0.125) | -0.316 (0.135) | -0.453 (0.121) |
| *FTH1* | Ferritin heavy chain | P02794 | 0.931 (0.247) | 0.914 (0.244) | 0.988 (0.249) | 0.726 (0.25) | 1.012 (0.282) | 0.931 (0.248) |
| *ARRB1* | Beta-arrestin-1 | P49407 | -0.386 (0.102) | -0.357 (0.101) | -0.388 (0.104) | -0.343 (0.106) | -0.317 (0.117) | -0.383 (0.102) |

**Table S5: P-values of covariates in linear regression model with COPD and one covariate**

**(Protein ~ COPD + covariate + batch effects + SVs)**

| Gene Name | Protein Name | Uniprot ID | Age | Sex | BMI | Pack-years | Cancer |
|---|---|---|---|---|---|---|---|
| *AGRN* | Agrin | O00468 | 2.68E-01 | 7.85E-01 | 2.81E-01 | 4.84E-01 | 3.61E-04 |
| *ANXA2* | Annexin A2 | P07355 | 4.03E-01 | 9.97E-01 | 8.14E-01 | 2.80E-02 | 6.32E-02 |
| *GPRC5A* | Retinoic acid-induced protein 3 | Q8NFJ5 | 3.99E-01 | 3.34E-01 | 5.20E-01 | 5.71E-01 | 5.40E-02 |
| *PLLP* | Plasmolipin | Q9Y342 | 5.72E-01 | 5.48E-01 | 7.19E-01 | 4.82E-01 | 1.28E-02 |
| *OCLN* | Occludin | Q16625 | 3.66E-01 | 7.83E-01 | 6.72E-01 | 1.76E-01 | 4.23E-01 |
| *LDHA* | L-lactate dehydrogenase A chain | P00338 | 3.61E-01 | 2.90E-02 | 7.84E-01 | 8.77E-01 | 9.72E-01 |
| *CAVIN1* | Caveolae-associated protein 1 | Q6NZI2 | 8.28E-01 | 8.27E-01 | 3.35E-01 | 1.43E-01 | 1.36E-01 |
| *IL33* | Interleukin-33 | O95760 | 5.01E-01 | 8.07E-01 | 1.30E-01 | 6.60E-02 | 6.20E-01 |
| *EHD2* | EH domain-containing protein 2 | Q9NZN4 | 9.49E-01 | 3.23E-01 | 2.80E-01 | 7.74E-02 | 1.29E-01 |
| *S100A10* | Protein S100-A10 | P60903 | 6.23E-01 | 4.56E-01 | 8.35E-01 | 7.76E-03 | 4.89E-01 |
| *EHD3* | EH domain-containing protein 3 | Q9NZN3 | 7.37E-01 | 6.55E-01 | 2.28E-01 | 7.60E-01 | 7.77E-01 |
| *FTL* | Ferritin light chain | P02792 | 4.66E-01 | 8.89E-01 | 4.27E-01 | 2.51E-02 | 2.74E-01 |
| *TNS3* | Tensin-3 | Q68CZ2 | 6.37E-02 | 9.96E-02 | 2.41E-01 | 7.11E-01 | 5.78E-01 |

**Table S5: P-values of covariates in linear regression model with COPD and one covariate**

**(Protein ~ COPD + covariate + batch effects + SVs)**

**(Continued)**

| Gene Name | Protein Name | Uniprot ID | Age | Sex | BMI | Pack-years | Cancer |
|---|---|---|---|---|---|---|---|
| *SUSD2* | Sushi domain-containing protein 2 | Q9UGT4 | 5.62E-01 | 1.85E-01 | 1.36E-01 | 1.32E-01 | 1.87E-03 |
| *DNAH5* | Dynein heavy chain 5, axonemal | Q8TE73 | 8.03E-01 | 1.42E-01 | 4.02E-01 | 1.39E-01 | 2.04E-01 |
| *ESAM* | Endothelial cell-selective adhesion molecule | Q96AP7 | 4.43E-01 | 6.88E-01 | 2.08E-01 | 8.44E-01 | 9.44E-01 |
| *RASIP1* | Ras-interacting protein 1 | Q5U651 | 6.67E-01 | 6.51E-01 | 8.76E-01 | 4.29E-01 | 7.87E-01 |
| *SRSF6* | Serine/arginine-rich splicing factor 6 | Q13247 | 5.93E-01 | 1.24E-01 | 5.08E-01 | 1.83E-01 | 8.22E-02 |
| *CAV1* | Caveolin-1 | Q03135 | 5.35E-01 | 5.00E-01 | 4.36E-01 | 1.42E-01 | 5.18E-02 |
| *AQP1* | Aquaporin-1 | P29972 | 2.29E-02 | 6.97E-01 | 2.40E-01 | 4.94E-02 | 1.62E-01 |
| *H3C1* | Histone H3.1 | P68431 | 8.67E-01 | 6.08E-01 | 6.57E-01 | 5.15E-01 | 6.18E-01 |
| *SFTPB* | Pulmonary surfactant-associated protein B | P07988 | 8.95E-01 | 7.31E-01 | 7.74E-01 | 2.06E-01 | 5.32E-01 |
| *LAMA4* | Laminin subunit alpha-4 | Q16363 | 4.72E-01 | 3.39E-01 | 9.15E-01 | 9.47E-01 | 2.96E-02 |
| *FTH1* | Ferritin heavy chain | P02794 | 6.98E-01 | 9.87E-01 | 2.49E-01 | 1.53E-02 | 5.10E-01 |
| *ARRB1* | Beta-arrestin-1 | P49407 | 1.38E-01 | 3.87E-01 | 2.66E-01 | 5.96E-01 | 5.04E-01 |

**Table S6: P-values of top proteins associated with COPD in linear regression model with only one covariate**

**(Protein ~ covariate + batch effects + SVs)**

| Gene Name | Protein Name | Uniprot ID | Age | Sex | BMI | Pack-years | Cancer |
|---|---|---|---|---|---|---|---|
| *AGRN* | Agrin | O00468 | 7.49E-02 | 6.44E-01 | 9.04E-01 | 4.37E-01 | 1.10E-07 |
| *ANXA2* | Annexin A2 | P07355 | 1.33E-01 | 8.27E-01 | 2.73E-01 | 7.51E-04 | 1.45E-04 |
| *GPRC5A* | Retinoic acid-induced protein 3 | Q8NFJ5 | 9.43E-01 | 2.69E-01 | 8.27E-01 | 7.04E-02 | 1.82E-04 |
| *PLLP* | Plasmolipin | Q9Y342 | 2.13E-01 | 7.11E-01 | 2.56E-01 | 5.59E-02 | 2.32E-05 |
| *OCLN* | Occludin | Q16625 | 8.77E-01 | 6.47E-01 | 7.08E-01 | 1.34E-02 | 7.26E-03 |
| *LDHA* | L-lactate dehydrogenase A chain | P00338 | 1.12E-01 | 2.48E-02 | 5.78E-01 | 1.57E-01 | 3.54E-02 |
| *CAVIN1* | Caveolae-associated protein 1 | Q6NZI2 | 6.53E-01 | 6.78E-01 | 9.25E-01 | 9.94E-03 | 1.08E-03 |
| *IL33* | Interleukin-33 | O95760 | 9.62E-01 | 6.84E-01 | 5.75E-01 | 3.80E-03 | 1.60E-02 |
| *EHD2* | EH domain-containing protein 2 | Q9NZN4 | 5.42E-01 | 2.59E-01 | 8.29E-01 | 4.29E-03 | 9.56E-04 |
| *S100A10* | Protein S100-A10 | P60903 | 2.60E-01 | 3.76E-01 | 3.08E-01 | 2.16E-04 | 8.79E-03 |
| *EHD3* | EH domain-containing protein 3 | Q9NZN3 | 7.21E-01 | 5.54E-01 | 7.64E-01 | 1.39E-01 | 7.91E-02 |
| *FTL* | Ferritin light chain | P02792 | 9.92E-01 | 9.60E-01 | 9.94E-01 | 1.22E-03 | 2.94E-01 |
| *TNS3* | Tensin-3 | Q68CZ2 | 2.57E-01 | 8.46E-02 | 7.50E-02 | 4.46E-01 | 2.37E-02 |

**Table S6: P-values of top proteins associated with COPD in linear regression model with only one covariate**

**(Protein ~ covariate + batch effects + SVs)**

**(Continued)**

| Gene Name | Protein Name | Uniprot ID | Age | Sex | BMI | Pack-years | Cancer |
|---|---|---|---|---|---|---|---|
| *SUSD2* | Sushi domain-containing protein 2 | Q9UGT4 | 2.28E-01 | 1.50E-01 | 5.45E-01 | 9.55E-03 | 3.55E-06 |
| *DNAH5* | Dynein heavy chain 5, axonemal | Q8TE73 | 7.31E-01 | 1.14E-01 | 1.18E-01 | 1.14E-02 | 3.33E-03 |
| *ESAM* | Endothelial cell-selective adhesion molecule | Q96AP7 | 1.58E-01 | 5.82E-01 | 6.69E-01 | 1.66E-01 | 5.57E-02 |
| *RASIP1* | Ras-interacting protein 1 | Q5U651 | 2.95E-01 | 5.46E-01 | 3.44E-01 | 5.74E-02 | 3.12E-02 |
| *SRSF6* | Serine/arginine-rich splicing factor 6 | Q13247 | 8.57E-01 | 1.02E-01 | 9.30E-01 | 1.72E-02 | 7.55E-04 |
| *CAV1* | Caveolin-1 | Q03135 | 2.22E-01 | 4.05E-01 | 9.74E-01 | 1.11E-02 | 3.30E-04 |
| *AQP1* | Aquaporin-1 | P29972 | 4.95E-03 | 5.75E-01 | 7.56E-01 | 2.56E-03 | 1.45E-03 |
| *H3C1* | Histone H3.1 | P68431 | 4.29E-01 | 4.97E-01 | 7.49E-01 | 5.84E-01 | 2.11E-02 |
| *SFTPB* | Pulmonary surfactant-associated protein B | P07988 | 4.48E-01 | 8.95E-01 | 3.15E-01 | 2.25E-02 | 1.63E-02 |
| *LAMA4* | Laminin subunit alpha-4 | Q16363 | 1.92E-01 | 4.63E-01 | 4.13E-01 | 2.33E-01 | 2.04E-04 |
| *FTH1* | Ferritin heavy chain | P02794 | 8.00E-01 | 8.58E-01 | 7.10E-01 | 8.60E-04 | 2.07E-01 |
| *ARRB1* | Beta-arrestin-1 | P49407 | 4.00E-01 | 3.29E-01 | 7.06E-01 | 1.22E-01 | 2.40E-02 |

**Table S7: P-values and Beta coefficients (Standard Error) of Pack-years for Top proteins associated with COPD in linear regression models separately in cases and controls**

**(Protein ~ pack-years + batch effects + SVs)**

| Gene Name | Protein Name | Uniprot ID | Control P-value | Control Beta-coefficient (standard error) | Case P-value | Case Beta-coefficient (standard error) | All Subject P-value | All Subject Beta-coefficient (standard error) |
|---|---|---|---|---|---|---|---|---|
| *AGRN* | Agrin | O00468 | 7.48E-01 | -0.002 (0.007) | 1.85E-01 | 0.004 (0.003) | 4.37E-01 | -0.002 (0.003) |
| *ANXA2* | Annexin A2 | P07355 | 5.70E-02 | -0.005 (0.003) | 1.30E-01 | -0.003 (0.002) | 7.51E-04 | -0.005 (0.001) |
| *GPRC5A* | Retinoic acid-induced protein 3 | Q8NFJ5 | 3.92E-01 | -0.005 (0.006) | 7.30E-01 | -0.002 (0.005) | 7.04E-02 | -0.007 (0.004) |
| *PLLP* | Plasmolipin | Q9Y342 | 3.24E-01 | -0.009 (0.009) | 7.09E-01 | -0.002 (0.005) | 5.59E-02 | -0.008 (0.004) |
| *OCLN* | Occludin | Q16625 | 2.23E-01 | -0.011 (0.009) | 2.53E-01 | -0.007 (0.006) | 1.34E-02 | -0.012 (0.005) |
| *LDHA* | L-lactate dehydrogenase A chain | P00338 | 3.87E-01 | 0.002 (0.002) | 5.41E-01 | -0.001 (0.001) | 1.57E-01 | 0.001 (0.001) |
| *CAVIN1* | Caveolae-associated protein 1 | Q6NZI2 | 1.80E-01 | -0.005 (0.004) | 3.66E-01 | -0.002 (0.002) | 9.94E-03 | -0.005 (0.002) |
| *IL33* | Interleukin-33 | O95760 | 2.73E-02 | -0.021 (0.009) | 3.18E-01 | -0.004 (0.004) | 3.80E-03 | -0.011 (0.004) |
| *EHD2* | EH domain-containing protein 2 | Q9NZN4 | 1.82E-01 | -0.006 (0.004) | 2.37E-01 | -0.003 (0.002) | 4.29E-03 | -0.006 (0.002) |
| *S100A10* | Protein S100-A10 | P60903 | 4.37E-03 | -0.01 (0.003) | 1.04E-01 | -0.004 (0.002) | 2.16E-04 | -0.007 (0.002) |
| *EHD3* | EH domain-containing protein 3 | Q9NZN3 | 3.97E-02 | -0.021 (0.01) | 5.12E-01 | 0.003 (0.005) | 1.39E-01 | -0.007 (0.004) |
| *FTL* | Ferritin light chain | P02792 | 2.54E-01 | 0.014 (0.012) | 5.37E-02 | 0.011 (0.006) | 1.22E-03 | 0.016 (0.005) |
| *TNS3* | Tensin-3 | Q68CZ2 | 9.53E-01 | <0.001(0.006) | 8.12E-01 | 0.001 (0.003) | 4.46E-01 | -0.002 (0.003) |

**Table S7: P-values and Beta coefficients (Standard Error) of Pack-years for Top proteins associated with COPD in linear regression models separately in cases and controls**

**(Protein ~ pack-years + batch effects + SVs)**

**(Continued)**

| Gene Name | Protein Name | Uniprot ID | Control P-value | Control Beta-coefficient (standard error) | Case P-value | Case Beta-coefficient (standard error) | All Subject P-value | All Subject Beta-coefficient (standard error) |
|---|---|---|---|---|---|---|---|---|
| *SUSD2* | Sushi domain-containing protein 2 | Q9UGT4 | 4.64E-01 | -0.004 (0.006) | 2.44E-01 | -0.004 (0.003) | 9.55E-03 | -0.007 (0.003) |
| *DNAH5* | Dynein heavy chain 5, axonemal | Q8TE73 | 3.58E-02 | -0.005 (0.002) | 2.44E-01 | -0.002 (0.002) | 1.14E-02 | -0.003 (0.001) |
| *ESAM* | Endothelial cell-selective adhesion molecule | Q96AP7 | 5.63E-01 | -0.004 (0.007) | 8.36E-01 | 0.001 (0.006) | 1.66E-01 | -0.006 (0.005) |
| *RASIP1* | Ras-interacting protein 1 | Q5U651 | 3.34E-02 | -0.018 (0.008) | 9.41E-01 | < 0.001 (0.006) | 5.74E-02 | -0.009 (0.005) |
| *SRSF6* | Serine/arginine-rich splicing factor 6 | Q13247 | 5.74E-01 | -0.003 (0.006) | 1.72E-01 | -0.005 (0.003) | 1.72E-02 | -0.006 (0.003) |
| *CAV1* | Caveolin-1 | Q03135 | 1.04E-01 | -0.008 (0.005) | 4.57E-01 | -0.002 (0.003) | 1.11E-02 | -0.006 (0.002) |
| *AQP1* | Aquaporin-1 | P29972 | 6.98E-01 | -0.002 (0.005) | 8.62E-02 | -0.005 (0.003) | 2.56E-03 | -0.007 (0.002) |
| *H3C1* | Histone H3.1 | P68431 | 4.49E-01 | -0.007 (0.009) | 5.68E-01 | -0.003 (0.005) | 5.84E-01 | 0.002 (0.004) |
| *SFTPB* | Pulmonary surfactant-associated protein B | P07988 | 1.70E-01 | -0.012 (0.009) | 8.84E-02 | 0.007 (0.004) | 2.25E-02 | 0.009 (0.004) |
| *LAMA4* | Laminin subunit alpha-4 | Q16363 | 2.42E-01 | 0.006 (0.005) | 9.55E-01 | < 0.001 (0.003) | 2.33E-01 | -0.003 (0.002) |
| *FTH1* | Ferritin heavy chain | P02794 | 1.72E-01 | 0.017 (0.012) | 5.94E-02 | 0.009 (0.005) | 8.60E-04 | 0.015 (0.004) |
| *ARRB1* | Beta-arrestin-1 | P49407 | 2.89E-01 | -0.004 (0.003) | 6.36E-01 | -0.001 (0.002) | 1.22E-01 | -0.003 (0.002) |

**Table S8: P-values and Beta coefficients (Standard Error) of Lung Cancer for Top proteins associated with COPD in linear regression models separately in cases and controls**

**(Protein ~ cancer + batch effects + SVs)**

| Gene Name | Protein Name | Uniprot ID | Control P-value | Control Beta-coefficient (standard error) | Case P-value | Case Beta-coefficient (standard error) | All Subject P-value | All Subject Beta-coefficient (standard error) |
|---|---|---|---|---|---|---|---|---|
| *AGRN* | Agrin | O00468 | 1.44E-01 | 0.525 (0.352) | 5.35E-03 | 0.587 (0.206) | 1.10E-07 | 0.848 (0.151) |
| *ANXA2* | Annexin A2 | P07355 | 3.88E-01 | 0.137 (0.157) | 6.40E-02 | 0.198 (0.106) | 1.45E-04 | 0.288 (0.074) |
| *GPRC5A* | Retinoic acid-induced protein 3 | Q8NFJ5 | 5.10E-01 | -0.227 (0.341) | 2.63E-02 | 0.649 (0.287) | 1.82E-04 | 0.722 (0.188) |
| *PLLP* | Plasmolipin | Q9Y342 | 4.42E-01 | -0.389 (0.502) | 3.76E-03 | 0.88 (0.296) | 2.32E-05 | 0.94 (0.215) |
| *OCLN* | Occludin | Q16625 | 4.74E-02 | 0.939 (0.46) | 6.31E-01 | 0.197 (0.409) | 7.26E-03 | 0.718 (0.263) |
| *LDHA* | L-lactate dehydrogenase A chain | P00338 | 1.85E-02 | 0.307 (0.125) | 2.18E-01 | -0.09 (0.072) | 3.54E-02 | -0.115 (0.054) |
| *CAVIN1* | Caveolae-associated protein 1 | Q6NZI2 | 9.25E-01 | 0.021 (0.219) | 8.87E-02 | 0.237 (0.138) | 1.08E-03 | 0.321 (0.096) |
| *IL33* | Interleukin-33 | O95760 | 9.20E-01 | -0.053 (0.525) | 7.82E-01 | 0.075 (0.271) | 1.60E-02 | 0.497 (0.204) |
| *EHD2* | EH domain-containing protein 2 | Q9NZN4 | 7.09E-01 | 0.093 (0.247) | 1.54E-01 | 0.208 (0.145) | 9.56E-04 | 0.347 (0.103) |
| *S100A10* | Protein S100-A10 | P60903 | 8.67E-01 | -0.035 (0.208) | 2.46E-01 | 0.166 (0.142) | 8.79E-03 | 0.264 (0.099) |
| *EHD3* | EH domain-containing protein 3 | Q9NZN3 | 2.26E-01 | -0.682 (0.555) | 7.68E-01 | 0.095 (0.321) | 7.91E-02 | 0.426 (0.241) |
| *FTL* | Ferritin light chain | P02792 | 5.20E-01 | 0.437 (0.674) | 2.94E-01 | 0.391 (0.37) | 2.94E-01 | -0.291 (0.276) |
| *TNS3* | Tensin-3 | Q68CZ2 | 8.42E-01 | -0.065 (0.323) | 3.63E-01 | 0.196 (0.215) | 2.37E-02 | 0.337 (0.148) |

**Table S8: P-values and Beta coefficients (Standard Error) of Lung Cancer for Top proteins associated with COPD in linear regression models separately in cases and controls**

**(Protein ~ cancer + batch effects + SVs)**

**(Continued)**

| Gene Name | Protein Name | Uniprot ID | Control P-value | Control Beta-coefficient (standard error) | Case P-value | Case Beta-coefficient (standard error) | All Subject P-value | All Subject Beta-coefficient (standard error) |
|---|---|---|---|---|---|---|---|---|
| *SUSD2* | Sushi domain-containing protein 2 | Q9UGT4 | 7.69E-01 | 0.093 (0.314) | 2.51E-03 | 0.662 (0.213) | 3.55E-06 | 0.697 (0.144) |
| *DNAH5* | Dynein heavy chain 5, axonemal | Q8TE73 | 1.02E-03 | 0.431 (0.122) | 8.21E-01 | 0.023 (0.099) | 3.33E-03 | 0.202 (0.068) |
| *ESAM* | Endothelial cell-selective adhesion [illegible] | Q96AP7 | 3.11E-01 | -0.41 (0.399) | 9.56E-01 | 0.021 (0.389) | 5.57E-02 | 0.476 (0.247) |
| *RASIP1* | Ras-interacting protein 1 | Q5U651 | 7.75E-01 | -0.134 (0.466) | 5.66E-01 | 0.223 (0.388) | 3.12E-02 | 0.555 (0.255) |
| *SRSF6* | Serine/arginine-rich splicing factor 6 | Q13247 | 1.87E-02 | 0.702 (0.287) | 2.52E-01 | 0.253 (0.22) | 7.55E-04 | 0.499 (0.145) |
| *CAV1* | Caveolin-1 | Q03135 | 7.35E-01 | -0.097 (0.285) | 2.59E-02 | 0.387 (0.171) | 3.30E-04 | 0.444 (0.121) |
| *AQP1* | Aquaporin-1 | P29972 | 2.48E-01 | -0.308 (0.263) | 2.69E-01 | 0.201 (0.181) | 1.45E-03 | 0.412 (0.127) |
| *H3C1* | Histone H3.1 | P68431 | 6.64E-01 | 0.224 (0.512) | 5.10E-01 | -0.203 (0.307) | 2.11E-02 | -0.526 (0.226) |
| *SFTPB* | Pulmonary surfactant-associated protein B | P07988 | 2.95E-01 | -0.498 (0.469) | 7.40E-01 | 0.09 (0.27) | 1.63E-02 | -0.502 (0.207) |
| *LAMA4* | Laminin subunit alpha-4 | Q16363 | 1.86E-01 | 0.35 (0.26) | 1.24E-01 | 0.25 (0.161) | 2.04E-04 | 0.45 (0.118) |
| *FTH1* | Ferritin heavy chain | P02794 | 2.13E-01 | 0.82 (0.648) | 6.00E-01 | 0.169 (0.321) | 2.07E-01 | -0.319 (0.251) |
| *ARRB1* | Beta-arrestin-1 | P49407 | 7.50E-01 | 0.061 (0.192) | 4.87E-01 | 0.109 (0.156) | 2.40E-02 | 0.235 (0.103) |

***Linear regression analysis on proteomic datasets***

After data preprocessing, we applied linear regression analysis on the normalized and imputed datasets (with surrogate variables) using linear regression models to identify candidate protein biomarkers for COPD. The candidate protein biomarkers were assessed with adjustment for age, sex, batch effects, and surrogate variables, and multiple statistical testing was adjusted with false discovery rates (FDR). The formula for linear regression analysis is shown below:

$$protein \sim case\ (0/1) + age + sex + column(batch\ effects) + surrogate\ variables\ (SVs)$$

P-values were corrected for multiple testing with the Benjamini-Hochberg False Discovery Rate (FDR) method. FDR was controlled at 5%. The detailed protein name, UniProt ID, gene symbol, and FDR of all features with FDR < 0.1 are shown in **Supplemental Table S9**.

**Table S9: Proteins associated with COPD with FDR less than 0.1 based on linear regression analysis (with age, sex, batch effects, and surrogate variables as covariates)**

| Gene Name | Protein Name | UniProt ID | FDR | Beta-coefficient | Standard error |
|---|---|---|---|---|---|
| *AGRN* | Agrin | O00468 | 0.019 | -0.732 | 0.159 |
| *ANXA2* | Annexin A2 | P07355 | 0.019 | -0.339 | 0.074 |
| *GPRC5A* | Retinoic acid-induced protein 3 | Q8NFJ5 | 0.019 | -0.857 | 0.189 |
| *IL33* | Interleukin-33 | O95760 | 0.023 | -0.850 | 0.202 |
| *LDHA* | L-lactate dehydrogenase A chain | P00338 | 0.023 | 0.220 | 0.052 |
| *FTL* | Ferritin light chain | P02792 | 0.023 | 1.113 | 0.27 |
| *S100A10* | Protein S100-A10 | P60903 | 0.023 | -0.409 | 0.098 |
| *OCLN* | Occludin | Q16625 | 0.023 | -1.123 | 0.262 |
| *CAVIN1* | Caveolae-associated protein 1 | Q6NZI2 | 0.023 | -0.407 | 0.097 |
| *EHD3* | EH domain-containing protein 3 | Q9NZN3 | 0.023 | -0.982 | 0.236 |
| *EHD2* | EH domain-containing protein 2 | Q9NZN4 | 0.023 | -0.432 | 0.104 |
| *PLLP* | Plasmolipin | Q9Y342 | 0.023 | -0.949 | 0.22 |
| *TENS3* | Tensin-3 | Q68CZ2 | 0.035 | -0.578 | 0.145 |
| *DNAH5* | Dynein heavy chain 5, axonemal | Q8TE73 | 0.035 | -0.266 | 0.067 |
| *SUSD2* | Sushi domain-containing protein 2 | Q9UGT4 | 0.035 | -0.596 | 0.151 |
| *CAV1* | Caveolin-1 | Q03135 | 0.038 | -0.475 | 0.123 |
| *SRSF6* | Serine/arginine-rich splicing factor 6 | Q13247 | 0.038 | -0.568 | 0.146 |
| *RASIP1* | Ras-interacting protein 1 | Q5U651 | 0.038 | -0.983 | 0.252 |
| *ESAM* | Endothelial cell-selective adhesion molecule | Q96AP7 | 0.038 | -0.953 | 0.244 |
| *AQP1* | Aquaporin-1 | P29972 | 0.045 | -0.476 | 0.125 |
| *H3C1* | Histone H3.1 | P68431 | 0.045 | 0.854 | 0.225 |
| *FTH1* | Ferritin heavy chain | P02794 | 0.047 | 0.931 | 0.248 |
| *SFTPB* | Pulmonary surfactant-associated protein B | P07988 | 0.047 | 0.780 | 0.207 |

**Table S9: Proteins associated with COPD with FDR less than 0.1 based on linear regression analysis (with age, sex, batch effects, and surrogate variables as covariates) (continued)**

| Gene Name | Protein Name | UniProt ID | FDR | Beta-coefficient | Standard error |
|---|---|---|---|---|---|
| *ARRB1* | arrestin beta 1 | P49407 | 0.047 | -0.383 | 0.102 |
| *LAMA4* | laminin subunit alpha 4 | Q16363 | 0.047 | -0.453 | 0.121 |
| *FOLR1* | folate receptor alpha | P15328 | 0.051 | -0.917 | 0.247 |
| *RPL35* | ribosomal protein L35 | P42766 | 0.056 | 0.439 | 0.12 |
| *NID1* | nidogen 1 | P14543 | 0.057 | -0.361 | 0.099 |
| *RALA* | RAS like proto-oncogene A | P11233 | 0.061 | -0.986 | 0.272 |
| *LAMC1* | laminin subunit gamma 1 | P11047 | 0.063 | -0.278 | 0.077 |
| *AGER* | advanced glycosylation end-product specific receptor | Q15109 | 0.063 | -0.676 | 0.188 |
| *PGD* | phosphogluconate dehydrogenase | P52209 | 0.071 | 0.266 | 0.075 |
| *AHNAK* | AHNAK nucleoprotein | Q09666 | 0.071 | -0.199 | 0.056 |
| *AKR1B1* | aldo-keto reductase family 1 member B | P15121 | 0.077 | 0.34 | 0.097 |
| *BPIFA1* | BPI fold containing family A member 1 | Q9NP55 | 0.077 | 0.458 | 0.131 |
| *TGM2* | transglutaminase 2 | P21980 | 0.079 | 0.284 | 0.081 |
| *HOXA5* | homeobox A5 | P20719 | 0.088 | -0.502 | 0.145 |
| *H6PD* | hexose-6-phosphate dehydrogenase/glucose 1-dehydrogenase | O95479 | 0.091 | 0.569 | 0.166 |
| *ATP1A1* | ATPase Na+/K+ transporting subunit alpha 1 | P05023 | 0.091 | -0.236 | 0.069 |
| *MZB1* | marginal zone B and B1 cell specific protein | Q8WU39 | 0.091 | 1.192 | 0.348 |
| *SH3BP1* | SH3 domain binding protein 1 | Q9Y3L3 | 0.091 | 1.038 | 0.304 |
| *CA3* | carbonic anhydrase 3 | P07451 | 0.094 | 0.642 | 0.189 |
| *RPS8* | ribosomal protein S8 | P62241 | 0.094 | 0.44 | 0.13 |
| *SOD2* | superoxide dismutase 2 | P04179 | 0.096 | 0.398 | 0.118 |
| *C5* | complement C5 | P01031 | 0.1 | 0.405 | 0.121 |

***Correlation analysis on proteomics datasets***

Based on the results of linear regression analysis, we chose the top 5% of proteins (n=220) according to FDR for Pearson correlation analysis (Benesty, Chen, Huang, & Cohen, 2009) on residualized values after removing the effects of age, sex, batch effects, and surrogate variables. To reveal the correlations between any two features (protein residuals), we applied R function *rcorr* in the *Hmisc* (version 4.4-0) package to compute Pearson correlation coefficients and statistical significance levels. Feature (protein) pairs with Pearson's correlation coefficients > 0.8 are shown in **Table 3**. We also constructed a scatter diagram to comprehensively show the correlations between the top correlated proteins (using residuals removing effects of age, sex, batch effects (columns) and surrogate variables) (**Supplemental Figure S4**). The comprehensive distribution pattern of the correlations between the top 5% of proteins, shown in **Supplemental Figure S5**, indicates that relatively few pairs of proteins had correlation coefficients above 0.8.

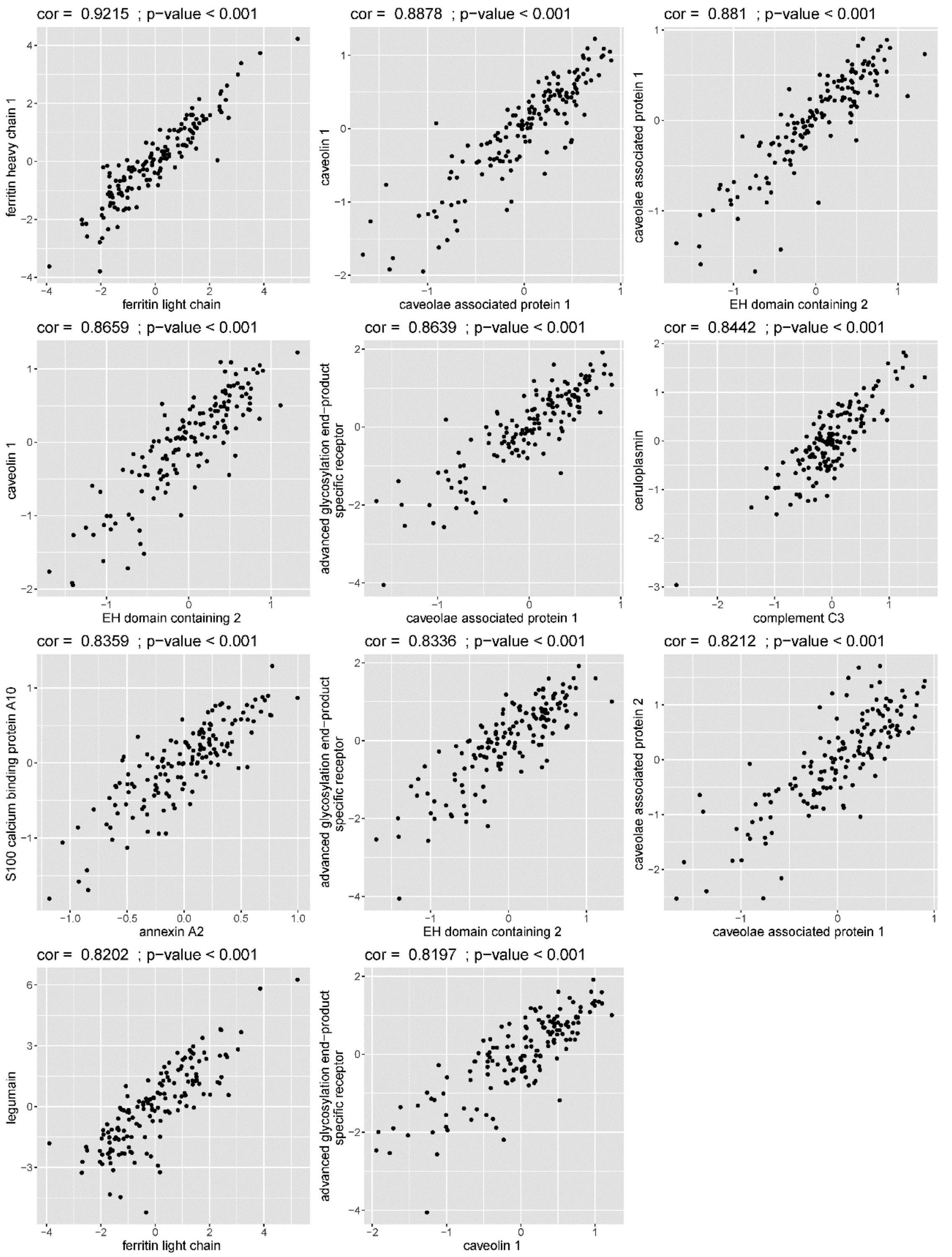


**Figure S4: Correlation scatter diagrams of top highly correlated proteins**. Pearson correlation values for protein residuals (removing effects of age, sex, batch effects (columns) and surrogate variables) between highly correlated proteins are shown.

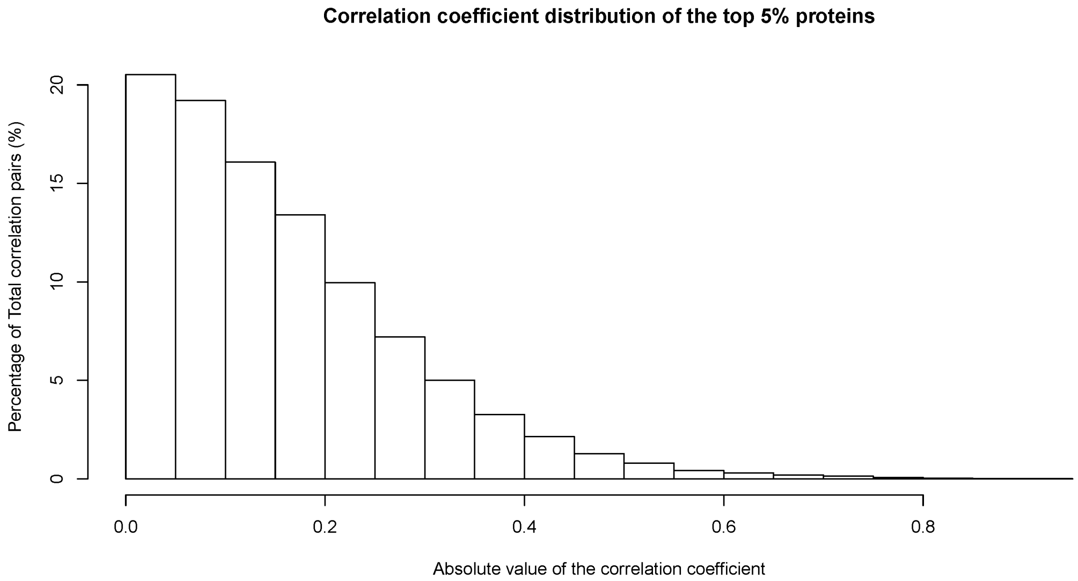


**Figure S5: Correlation coefficient distribution of the top 5% proteins.** Residual values (removing effects of age, sex, batch effects (columns) and surrogate variables) were used to calculate the correlations between the top 5% of proteins. In this histogram, we showed the distribution of the absolute value of the correlation coefficient between each pair of the top 5% proteins. Most pairs of proteins have relatively low correlation coefficients. However, a small number of highly correlated proteins were found in the top 5% of proteins.

### *Machine Learning analysis on proteomics datasets*

Using the top 5% of proteins (n=220) identified from the single protein regression models, we performed machine learning analysis on proteomics residuals removing age, sex, column (batch effects), and surrogate variables. Since our dataset is relatively small and did not include a replication dataset, we applied 5-fold cross-validation using three widely used machine learning models (Random Forest, Elastic Net, and Naïve Bayes). The averaged accuracy, AUC, and feature importance were compared. To perform feature selection inside the cross-validation, we applied linear regression analysis with training sets and identified top 220 features as candidates for further machine learning analysis.

The first method used was Random Forest. We used R function *train* in package *caret* (version 6.6-

86) (methods as “rf”) and R package *randomForest* (version 4.6-14)(Breiman, 2001)with no resampling approach with ntrees and mtry tuning to find the best parameters for Random Forest in each round of cross-validation. For each round of cross-validation, we calculated the accuracy and AUC to reflect the performance of the model. In addition to Random Forest, we also applied Elastic Net (Elastic net model paths for some generalized linear models) and Naïve Bayes for comparison. We built these models using R function *train* in package *caret* (version 6.6-86) with methods as “glmnet” and “nb”, respectively. Those methods were performed using R package *glmnet* (version 4.0-2) (Elastic Net) (Friedman, Hastie, & Tibshirani, 2009) and *klaR* (version 0.6-15)(Garczarek, 2002) (Naïve Bayes). For Elastic Net models, alpha and lambda were chosen as tuning parameters to build the optimal model inside the cross validation, while FL and adjust were chosen as tuning parameters for the Naïve Bayes model to optimize our machine learning models. The detailed workflow for feature selection and cross-validation resampling processes can be seen in **Supplemental Figure S1**.

We assessed feature importance for Random Forests using R function *varImp* from the *caret* package (Kuhn, 2008). For the entire cross-validation procedure, the final accuracy, AUC, and feature importance are all the averages of each round.

***Gene ontology enrichment analysis***

To investigate the potential biological functions of our COPD-associated protein biomarkers, gene ontology enrichment analysis was applied to to three groups of COPD-associated proteins identified from the linear regression models: the top 5% of proteins identified from the linear regression models (candidate protein biomarkers), COPD positively correlated proteins among the top 5% proteins (with positive beta coefficients) and COPD negatively correlated proteins among the top 5% proteins (with negative beta coefficients) using the R package *topGO* (version 2.38.1)(Alexa, Rahnenführer, & Lengauer, 2006). There are multiple algorithms provided by *topGO* that can be applied to identify statistically significant GO terms. *Elim* and *weight* algorithms are two widely used algorithms for GO enrichment analysis (Alexa et al., 2006). The *Elim* algorithm repeatedly removes genes that can be mapped to significant GO terms from high level (general) GO terms, while the *weight* algorithm identifies and annotates genes to specific GO terms based on the score of its neighboring GO terms. As the default algorithm of *topGO*, the weight01 algorithm combined the *removal* method (*Elim*) and *identification* method (*weight*). Therefore, to adjust for multiple comparisons and obtain a comprehensive enrichment result for our identified protein biomarkers, we used the “weight01” algorithm to identify significant GO terms. The threshold for the weight01 score was set as 0.001. The background set of genes included all of the proteins remaining after data preprocessing (4407 proteins). The enriched GO terms of top 5% proteins are shown in **Supplemental Table S10.** The enriched GO terms of COPD positively and negatively correlated proteins among the top 5% proteins with weight01 score < 0.001 can be seen in **Supplemental Figure S6 and S7**. The detailed enrichment results of COPD positively and negatively correlated proteins among the top 5% proteins are shown in **Supplemental Table S11 and S12**.

**Table S10: Gene ontology enrichment analysis on candidate protein biomarkers for COPD (top 5% of proteins)**

| GO.ID | Term | Annotated | Significant | Expected | Rank in classicFisher | classicFisher | Elim Fisher | Topgo Fisher | Parentchild Fisher | **cluster** |
|---|---|---|---|---|---|---|---|---|---|---|
| GO:0044331 | cell-cell adhesion mediated by cadherin | 15 | 6 | 0.75 | 30 | 5.100E-05 | 5.20E-05 | 1.90E-04 | 0.0019 | BP |
| GO:0007156 | homophilic cell adhesion via plasma membrane adhesion molecules | 20 | 6 | 1.01 | 51 | 0.00032 | 0.00032 | 3.20E-04 | 0.3621 | BP |
| GO:0071280 | cellular response to copper ion | 4 | 3 | 0.20 | 56 | 0.00048 | 0.00048 | 4.80E-04 | 0.0018 | BP |
| GO:0016323 | basolateral plasma membrane | 79 | 15 | 3.95 | 5 | 5.90E-06 | 5.90E-06 | 8.0E-06 | 0.03265 | CC |
| GO:0070062 | extracellular exosome | 1308 | 93 | 65.48 | 13 | 3.20E-05 | 3.20E-05 | 3.20E-05 | 0.34530 | CC |
| GO:0005923 | bicellular tight junction | 46 | 10 | 2.30 | 18 | 6.70E-05 | 6.70E-05 | 6.70E-05 | 0.20546 | CC |
| GO:0016328 | lateral plasma membrane | 24 | 7 | 1.20 | 22 | 0.00012 | 0.00012 | 1.20E-04 | 0.00481 | CC |
| GO:0042383 | sarcolemma | 59 | 13 | 2.95 | 4 | 4.50E-06 | 0.00019 | 1.90E-04 | 0.00013 | CC |
| GO:0030659 | cytoplasmic vesicle membrane | 337 | 22 | 16.87 | 200 | 0.11635 | 0.11635 | 2.60E-04 | 0.55954 | CC |
| GO:0005788 | endoplasmic reticulum lumen | 151 | 18 | 7.56 | 31 | 0.00045 | 0.00045 | 4.50E-04 | 0.00060 | CC |
| GO:0019834 | phospholipase A2 inhibitor activity | 4 | 3 | 0.20 | 3 | 0.00047 | 0.00047 | 4.70E-04 | 0.11 | MF |

[a] Fisher-test of weight01 algorithm: P-value < 0.001

**Table S11: Gene ontology enrichment analysis on candidate protein biomarkers for COPD**

**(COPD positively associated proteins in the top 5% proteins)**

| GO.ID | Term | Annotated | Significant | Expected | Rank in classicFisher | Weight01 Fisher[a] | classicFisher | Elim Fisher | Parentchild Fisher | cluster |
|---|---|---|---|---|---|---|---|---|---|---|
| GO:0043312 | neutrophil degranulation | 354 | 18 | 7.64 | 23 | 4.40E-04 | 0.00044 | 0.00044 | 0.36 | BP |
| GO:0006006 | glucose metabolic process | 86 | 8 | 1.86 | 25 | 6.80E-04 | 0.00046 | 0.01703 | 0.57 | BP |
| GO:0004322 | ferroxidase activity | 2 | 2 | 0.04 | 1 | 4.40E-04 | 0.00044 | 0.00044 | 1 | MF |
| GO:0070062 | extracellular exosome | 1308 | 50 | 28.02 | 4 | 1.00E-06 | 1.20E-06 | 1.20E-06 | 0.56442 | CC |
| GO:1904724 | tertiary granule lumen | 44 | 6 | 0.94 | 11 | 3.00E-04 | 0.0003 | 0.0003 | 0.00113 | CC |
| GO:0008043 | intracellular ferritin complex | 2 | 2 | 0.04 | 16 | 4.50E-04 | 0.00045 | 0.00045 | 0.00044 | CC |
| GO:0034663 | endoplasmic reticulum chaperone complex | 9 | 3 | 0.19 | 18 | 7.30E-04 | 0.00073 | 0.00073 | 0.00081 | CC |

[a] Fisher-test of weight01 algorithm: P-value < 0.001

**Table S12: Gene ontology enrichment analysis on candidate protein biomarkers for COPD**

**(COPD negatively associated proteins in the top 5% proteins)**

| GO.ID | Term | Annotated | Significant | Expected | Rank in classicFisher | Weight01 Fisher[a] | classicFisher | Elim Fisher | Parentchild Fisher | cluster |
|---|---|---|---|---|---|---|---|---|---|---|
| GO:0007156 | homophilic cell adhesion via plasma membrane adhesion molecules | 20 | 6 | 0.57 | 33 | 1.40E-05 | 1.40E-05 | 1.40E-05 | 0.24494 | BP |
| GO:0044331 | cell-cell adhesion mediated by cadherin | 15 | 6 | 0.43 | 15 | 2.00E-05 | 2.00E-06 | 2.00E-06 | 0.00018 | BP |
| GO:0034332 | adherens junction organization | 72 | 9 | 2.07 | 59 | 7.60E-05 | 0.00018 | 0.00018 | 0.45625 | BP |
| GO:0072659 | protein localization to plasma membrane | 122 | 11 | 3.5 | 81 | 1.70E-04 | 0.00066 | 0.00066 | 0.04854 | BP |
| GO:0016339 | calcium-dependent cell-cell adhesion via plasma membrane cell adhesion molecules | 6 | 3 | 0.17 | 76 | 4.30E-04 | 0.00043 | 0.00043 | 0.11966 | BP |
| GO:0007010 | cytoskeleton organization | 514 | 34 | 14.76 | 12 | 7.20E-04 | 1.40E-06 | 0.00069 | 9.30E-06 | BP |
| GO:0005509 | calcium ion binding | 210 | 17 | 6.04 | 1 | 8.50E-05 | 8.50E-05 | 8.50E-05 | 7.90E-05 | MF |
| GO:0019834 | phospholipase A2 inhibitor activity | 4 | 3 | 0.12 | 2 | 9.10E-05 | 9.10E-05 | 9.10E-05 | 0.1143 | MF |
| GO:0005544 | calcium-dependent phospholipid binding | 15 | 4 | 0.43 | 5 | 7.00E-04 | 7.00E-04 | 7.00E-04 | 0.0028 | MF |
| GO:0016323 | basolateral plasma membrane | 79 | 13 | 2.26 | 7 | 1.00E-06 | 2.50E-07 | 2.50E-07 | 0.03073 | CC |

[a] Fisher-test of weight01 algorithm: P-value < 0.001

**Table S12: Gene ontology enrichment analysis on candidate protein biomarkers for COPD**

**(COPD negatively associated proteins in the top 5% proteins, continued)**

| GO.ID | Term | Annotated | Significant | Expected | Rank in classicFisher | Weight01 Fisher[a] | classicFisher | Elim Fisher | Parentchild Fisher | cluster |
|---|---|---|---|---|---|---|---|---|---|---|
| GO:0016328 | lateral plasma membrane | 24 | 7 | 0.69 | 11 | 3.00E-06 | 3.10E-06 | 3.10E-06 | 0.00083 | CC |
| GO:0030659 | cytoplasmic vesicle membrane | 337 | 18 | 9.65 | 56 | 4.00E-06 | 0.00681 | 0.00681 | 0.00756 | CC |
| GO:0042383 | sarcolemma | 59 | 12 | 1.69 | 6 | 1.80E-05 | 6.40E-08 | 1.80E-05 | 1.90E-05 | CC |
| GO:0005923 | bicellular tight junction | 46 | 8 | 1.32 | 19 | 3.80E-05 | 3.80E-05 | 3.80E-05 | 0.32163 | CC |
| GO:0016324 | apical plasma membrane | 97 | 10 | 2.78 | 28 | 3.90E-04 | 0.00039 | 0.00039 | 0.53642 | CC |
| GO:0005901 | caveola | 32 | 8 | 0.92 | 9 | 4.40E-04 | 2.10E-06 | 0.00015 | 0.02787 | CC |
| GO:0002095 | caveolar macromolecular signaling complex | 2 | 2 | 0.06 | 32 | 8.10E-04 | 0.00081 | 0.00081 | 0.0043 | CC |
| GO:1990665 | AnxA2-p11 complex | 2 | 2 | 0.06 | 33 | 8.10E-04 | 0.00081 | 0.00081 | 0.001 | CC |
| GO:1990812 | growth cone filopodium | 2 | 2 | 0.06 | 34 | 8.10E-04 | 0.00081 | 0.00081 | 0.00209 | CC |
| GO:0030315 | T-tubule | 16 | 4 | 0.46 | 35 | 8.90E-04 | 0.00089 | 0.00089 | 0.0222 | CC |

[a] Fisher-test of weight01 algorithm: P-value < 0.001

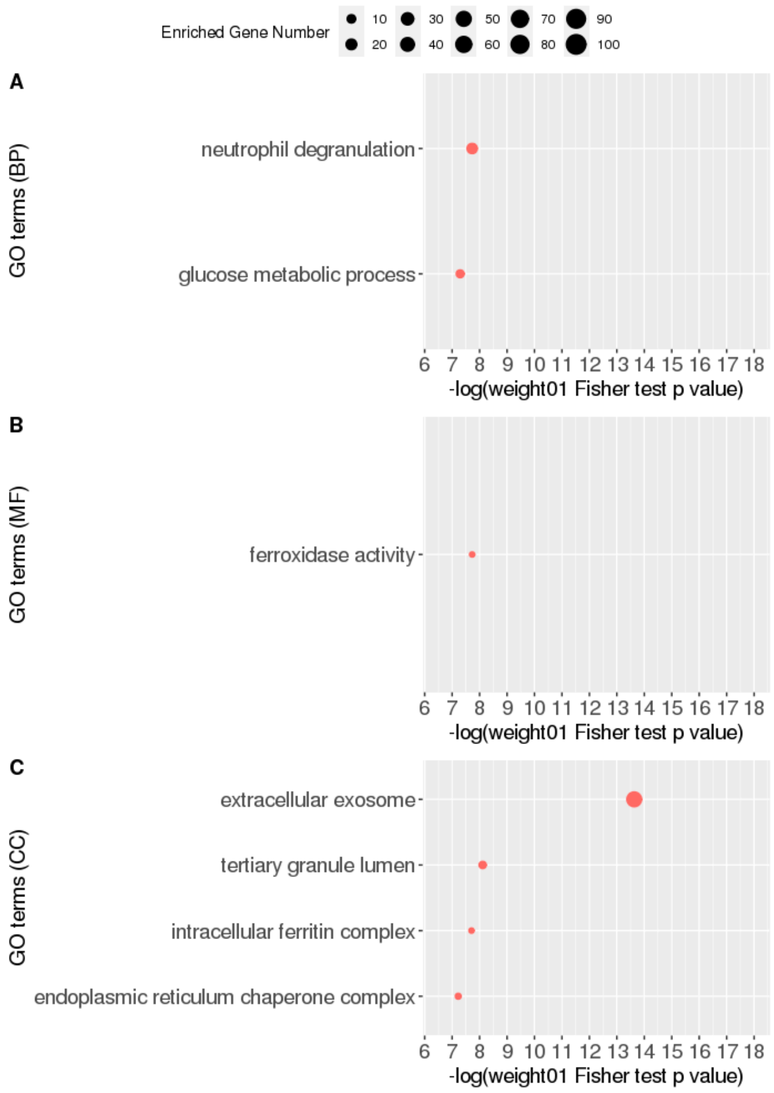


**Figure S6: Gene ontology enrichment analysis on candidate protein biomarkers for COPD (top 5% of COPD positively correlated proteins according to FDR in linear regression analysis).** Gene ontology enrichment analysis was performed on candidate protein biomarkers at three levels: A) biological processes (BP); B) molecular functions (MF); and C) cellular components (CC) with weight01 score threshold. The x-axis indicates the negative log value of weight01 Fisher test p-value. The more significantly the GO term enriched, the higher the value on the x-axis. The y-axis indicates different GO terms ranked by the weight01 Fisher test p-value. The size of the point in the figure indicates the number of genes significantly enriched in this GO term.

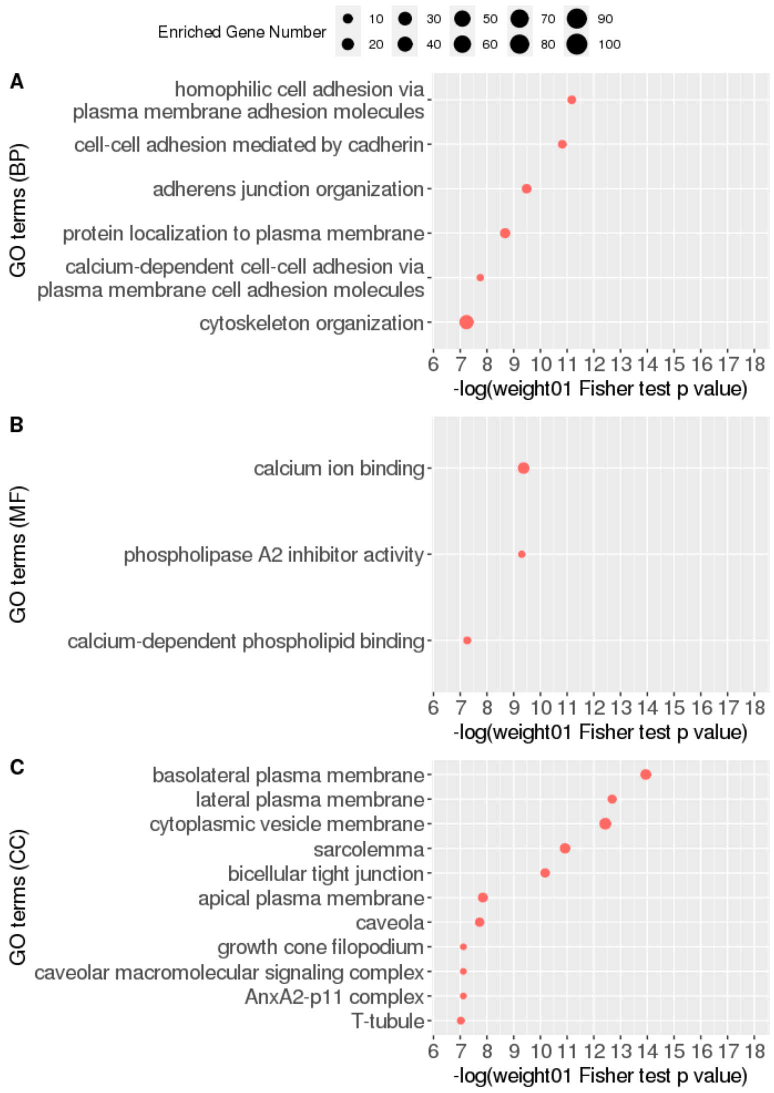


**Figure S7: Gene ontology enrichment analysis on candidate protein biomarkers for COPD (top 5% of COPD negatively correlated proteins according to FDR in linear regression analysis).** Gene ontology enrichment analysis was performed on candidate protein biomarkers at three levels: A) biological processes (BP); B) molecular functions (MF); and C) cellular components (CC) with weight01 score threshold. The x-axis indicates the negative log value of weight01 Fisher test p-value. The more significantly the GO term enriched, the higher the value on the x-axis. The y-axis indicates different GO terms ranked by the weight01 Fisher test p-value. The size of the point in the figure indicates the number of genes significantly enriched in this GO term.
